# Addressing challenges in selecting dietary risk factor-outcome pairs for health impact assessments: Developing an evidence database and novel approach to dose-response extraction

**DOI:** 10.1101/2025.08.27.25333764

**Authors:** Anna E. Jacob, Claire Denos, Lieselot Boone, Margot Cooreman-Algoed, Jo Dewulf, Marianne Uhre Jakobsen, Stefanie Vandevijvere, Sara M. Pires, Lea Sletting Jakobsen

## Abstract

Dietary health impact assessments are valuable decision-support tools for action on population-wide dietary shifts. These assessments rely on epidemiological evidence to estimate associations between dietary risk factors and health outcomes. However, the review and selection of evidence poses several challenges including a lack of systematic approaches to risk-outcome pair selection and ability to extract harmonised dose-response data from scientific papers. To address these challenges, this work aimed to 1) develop a database of dietary risk factor dose-response curves by updating the Nordic Nutrition Recommendations 2023 (NNR2023) evidence base and 2) propose a novel extraction method for non-linear dose-response curves. The NNR2023 scoping review methodology was replicated and risk-outcome pairs were selected according to modified NNR 2023 criteria. Non-linear relationships were estimated by extracting data points with an open-source graph reader and fitting piecewise constant functions to the extracted points. From the selected updated studies and original NNR2023 evidence, 101 risk-outcome associations were included, of which 44 were non-linear, and one quarter included beverages. Around half of the associations reported levels of at least mid-range evidence certainty. The provided database and methodology contribute to increased transparency and provide a standard approach to evidence selection and use in health impact assessments.

## Introduction

Health impact assessments, including dietary risk factor modelling, optimisations, and comparative risk assessments, are valuable decision support tools that are growing in popularity in nutritional and public health related fields [1,2]. These approaches estimate how health (and other relevant) impacts of hypothetical counterfactual dietary scenarios could affect a population. However, they often require researchers to rely on published epidemiological risk data to form conclusions. This introduces an array of complex challenges that are seemingly rarely addressed in scientific literature but hamper interpretation across studies.

Risk estimates, including relative risk (RR), risk ratio, odds ratio, and hazard rate (referred to as RR for simplicity) are epidemiological measures that indicate the likelihood/rate/or odds of health outcomes in a population. Selection of risk-outcome pairs in dietary modelling already poses a challenge, but the dose-response functions for these pairs are also not readily available in many meta-analytic studies, nor in the large compilations of evidence. Use of the provided RR values in assessments requires modellers to either carefully reproduce meta-analytic results — which poses a major challenge for models integrating numerous risk-outcome pairs such as whole diet assessments — or limit risk-outcome pair selection to easily replicable linear associations, which may not accurately reflect the nature of the association. The evident lack of standardisation when it comes to dietary risk factor evidence selection can result in cherry-picking or relying on a single source of evidence, thereby impacting the bias of resulting assessments.

A recent scoping review highlighted the lack of alignment in data feeding into various types of dietary assessment models, with 44% of modelling studies using GBD data, while other studies used selected systematic reviews or other sources [2]. Although attempts to summarise and/or standardise these data sources and related review methodologies have been made (the Global Burden of Disease (GBD) study, World Cancer Research Fund (WCRF), and Nordic Nutrition Recommendations (NNR)) [3–5], there are limitations that must be considered and transparently communicated in the works that subsequently use these data.

The GBD, WCRF, and NNR all consisted of large teams of experts conducting in-depth reviews of dietary evidence, each with its own set of strengths and weaknesses and assessment criteria. Following many improvements to earlier renditions, the 2021 GBD study included detailed non-linear dose-response information for dietary risk factors [3,6]. While the GBD’s efforts are generally considered to be reliable and high-quality, critiques also exist [7–10]. The transparency of the GBD methodology has been an ongoing challenge for many. For example, results from only two dietary risk factors (red meat and vegetables) had been published in peer-reviewed formats at the time of our update, with processed meat and sugar-sweetened beverages (SSBs) published after [6,11–13]. The WCRF also makes continuous efforts to quantify the risk of cancer associated with dietary intakes through the Global Cancer Update Program [4]. This project is thoroughly conducted, and results are well-organised and presented, but outcomes are limited to cancer and similar issues of data extraction with GBD, particularly for non-linear associations arise. Both initiatives are valuable sources of data and commendable for their efforts in the field; however, NNR2023 appears to cover the most breadth as an evidence base, including both GBD and WCRF results within its own scope.

The NNR2023 was commissioned by the Nordic Council of Ministers and included a list of “qualified systematic reviews” for each food category of interest to provide causal scientific evidence for the recommendations [5]. While the period for evidence inclusion varied slightly between food groups of interest, the general timeframe was approximately ten years prior to NNR2023 search dates. Unlike the GBD and WCRF, NNR2023 did not conduct meta-analyses from their selected evidence but rather used the data as the justification for various dietary guidelines. The methodology for selection and development of the so-called qualified systematic reviews along with the methodology for conducting scoping reviews for each food group were rigid, transparent, and recorded in a series of background papers [5,14–29]. Due to the replicability and thoroughness in reviewing relevant dietary data, including both the WCRF and peer-reviewed GBD results in its scope, NNR2023 was determined to have the most robust evidence base of the three sources. The pace and volume of data being published regarding dietary health risks presents a formidable challenge to staying current with the evidence. We therefore decided to conduct an update of the relevant scoping review searches prior to selecting risk-outcome pairs, not only to identify any new systematic reviews to include since the last search date, but to elucidate the challenge of maintaining currency with the latest research.

This paper aimed to address challenges in data selection for dietary health impact modelling by 1) constructing an open-access database of dietary risk-outcome pairs and their accompanying dose-response associations using NNR2023 as the backbone evidence base, supplemented with an updated search and 2) proposing a novel and efficient method of extracting dose-response curves from such evidence sources.

## Methods

### Updating the NNR Qualified Systematic Reviews

The qualified systematic reviews listed in NNR2023 for food groups were investigated for RR estimates represented by quantitative dose-response data. These reviews were screened in duplicate, and the excluded studies were recorded (Appendix A, Table A1). Those that contained dose-response data were included.

An updated search was subsequently conducted to account for systematic reviews published after April 15^th^, 2023, which was the last search date for NNR2023 [5]. The search was organised for each of the food groups in NNR2023 according to the methodology of their respective scoping reviews and the NNR2023 methodology papers (i.e. beverages; cereals (grains); vegetables, fruits and berries; potatoes; fruit juices; pulses (legumes); nuts and seeds; fish and seafood; red meat; white meat; milk and dairy products; eggs; fats & oils; sweets; and alcohol) [5,15–19,21–26,30]. Detailed food group definitions can be found in Appendix B and alignment of food items and groups between NNR2023 and GBD can be found in Table B1. The search strings from the scoping review background papers were modified when needed (due to missing structure or details for searching databases) and implemented in the corresponding database(s), most often PubMed (Appendix C, Table C1). The search was conducted on August 2^nd^, 2024 (September 5^th^, 2024, for the Coffee and Tea group).

Identified sources were uploaded to Rayyan, a review screening software, where the articles were screened by a single reviewer against the modified NNR qualified systematic review selection criteria [31]. Three modifications were made by current review authors for thoroughness: (1) the additional condition of requiring dose-response data, (2) omitting the criterion of needing to be commissioned by national food or health authorities, or international food and health organisations, and (3) considering only foods or food groups as our scope did not include nutrient investigations (Table 1) [14,31].

**Table 1.**
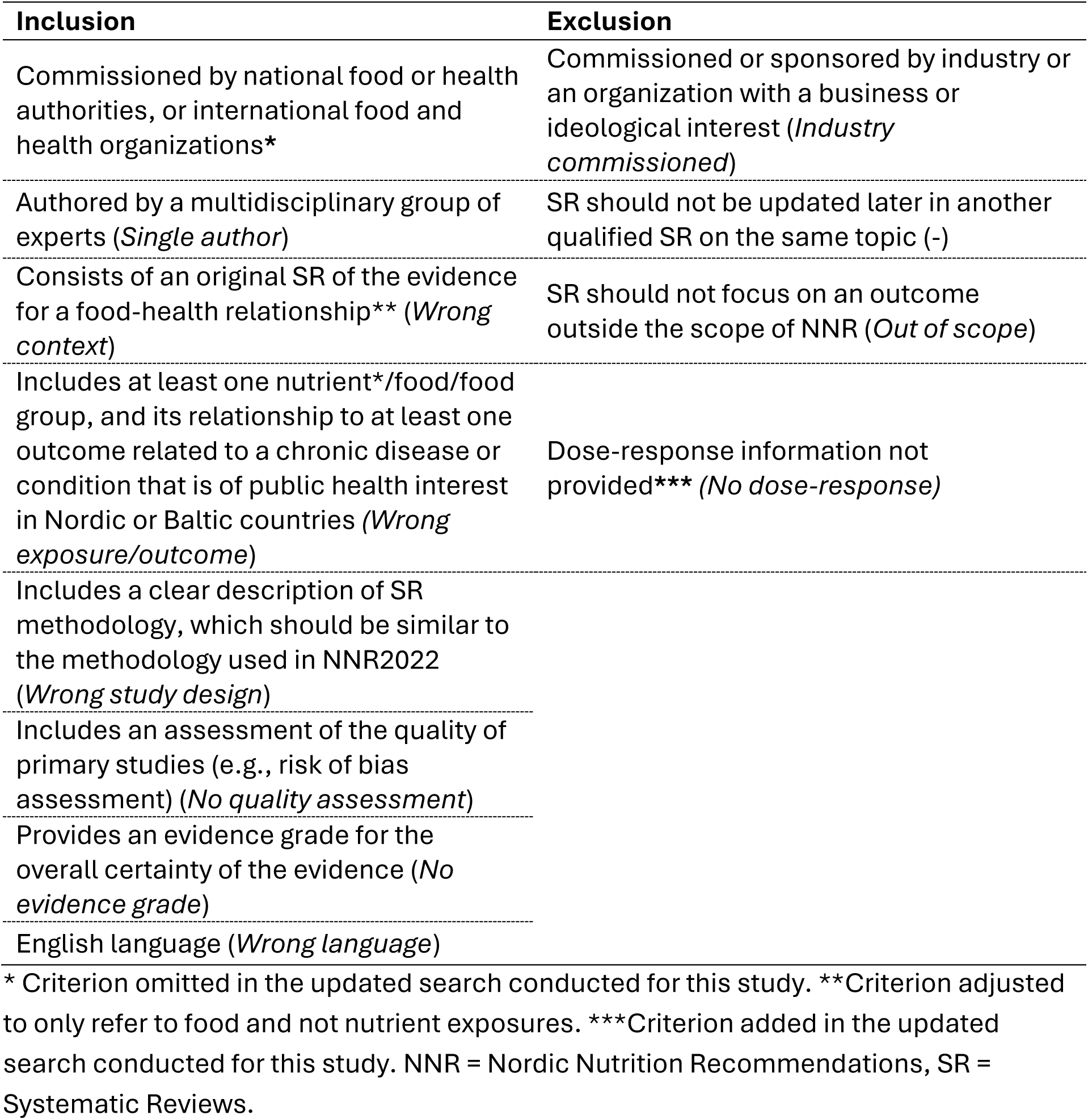
Selection criteria for systematic reviews found in the updated search, adapted from NNR2023 selection criteria for qualified systematic reviews [14]. *(Labels as used in the flow chart (Figure 1))*.

| Inclusion | Exclusion |
| --- | --- |
| Commissioned by national food or health authorities, or international food and health organizations* | Commissioned or sponsored by industry or an organization with a business or ideological interest (*Industry commissioned*) |
| Authored by a multidisciplinary group of experts (*Single author*) | SR should not be updated later in another qualified SR on the same topic (-) |
| Consists of an original SR of the evidence for a food-health relationship** (*Wrong context*) | SR should not focus on an outcome outside the scope of NNR (*Out of scope*) |
| Includes at least one nutrient*/food/food group, and its relationship to at least one outcome related to a chronic disease or condition that is of public health interest in Nordic or Baltic countries (*Wrong exposure/outcome*) | Dose-response information not provided*** (*No dose-response*) |
| Includes a clear description of SR methodology, which should be similar to the methodology used in NNR2022 (*Wrong study design*) |  |
| Includes an assessment of the quality of primary studies (e.g., risk of bias assessment) (*No quality assessment*) |  |
| Provides an evidence grade for the overall certainty of the evidence (*No evidence grade*) |  |
| English language (*Wrong language*) |  |

\* Criterion omitted in the updated search conducted for this study. \*\*Criterion adjusted to only refer to food and not nutrient exposures. \*\*\*Criterion added in the updated search conducted for this study. NNR = Nordic Nutrition Recommendations, SR = Systematic Reviews.

### Selecting Risk-Outcome Pairs

Once a list of systematic reviews was compiled, both from the NNR2023 qualified systematic reviews and the updated search, individual risk-outcome pairs were screened and dose-response estimates extracted. For associations found in the updated search, this process was completed in duplicate and included information on exposure, outcome, dose, certainty of evidence (according to one of four systems encountered: GRADE (Grading of Recommendations, Assessment, Development, and Evaluations), NutriGrade, WCRF grading, or burden of proof star ratings), number of studies included, and the date of last search. Conflicts from duplicate screening were resolved through discussion between reviewers. The same process was conducted for included papers from NNR2023 but was completed by a single reviewer. Reasons for inclusion and exclusion were summarised in the flow chart.

Risk-outcome pairs were only included if they were statistically significant, included more than one study in the dose-response calculation, and the provided grade for overall certainty of evidence was a minimum of low (GRADE and NutriGrade), limited-suggestive (WCRF), or two or more stars (GBD) [32–34]. For an overview of the certainty grading systems encountered, the four scales were mapped in comparison to one another based on qualitative descriptions.

### Summarising the Evidence

Following screening and selection, any risk-outcome pairs that were conflicting (e.g. different meta-analysis for the same risk-outcome pair) underwent a critical side-by-side comparison of studies to determine which had more robust or recent data (Appendix D). The decision was based on the recency of the meta-analysis, number of included studies, overlap of included studies, additional statistical measures, and if the quality and robustness were indistinguishable, an AMSTAR-2 assessment would have been applied if all else was equal [35]. The risk-outcome pairs from studies that were deemed stronger in the comparisons were included.

### Extraction of Dose-Response Associations

#### Linear Associations

If the association was linear or decidedly linear (i.e. there was no evidence or investigation of non-linearity), the effect estimate (RR) and unit of exposure was extracted with any associated uncertainty (95% CI). In the building of the database, the slope of the function was calculated with the assumption that the relationship between exposure and RR follows a log-linear relationship according to Berlin et al. [36].

#### Non-Linear Associations

When an association was identified as non-linear, such as when a significant p-value for non-linearity was disclosed in the paper (i.e. p < 0.05), the following protocol was applied:

1. **Extract available curves:** If the underlying data from the meta-analyses (RR for incremental dose sections, etc.) or functions of the curves were included in the publication, these were extracted directly.
2. **Use a piecewise approach:** In the case the underlying data was not readily available or provided, an online AI graph reader (WebPlotDigitiser) was used with a piecewise linear approach to estimate the non-linear curves [37]. Points were extracted from a calibrated visual copy of the curve, and a function which applied short piecewise linear functions for each data point was applied. The assumption was made that the curve continued as levelled off from the last piece of the function. In the case datapoints did not start at zero, a similar assumption was applied, that all values between zero and the first datapoint were equal to the first datapoint. The detailed process for this procedure is explained in Appendix E.

For comparison and completeness, Burden of Proof outcomes that met the minimum evidence grade cut-off of two stars (and were not included in the NNR), were also extracted from the VizHub tool and summarised in Appendix F, Table F1 [6]. Alcohol is not included as a dietary risk factor in GBD, but we have included it here to align with NNR2023 food groups.

An open-access database of extracted dose-response curves for the selected risk-outcome pairs was constructed using RStudio [38]. This database is available on Zenodo [39].

## Results

In total, 101 risk-outcome pairs were selected from the NNR2023 qualified systematic reviews (n=66) and the updated search results (n=35). Of these, 44 followed non-linear associations (Table 2). The associations were reported by food groups (n=11) and individual food items. Notably, a quarter of the associations involved non-alcoholic beverages or alcohol and no studies on sweets (excluding SSBs), potatoes, or pulses were included.

**Table 2.**
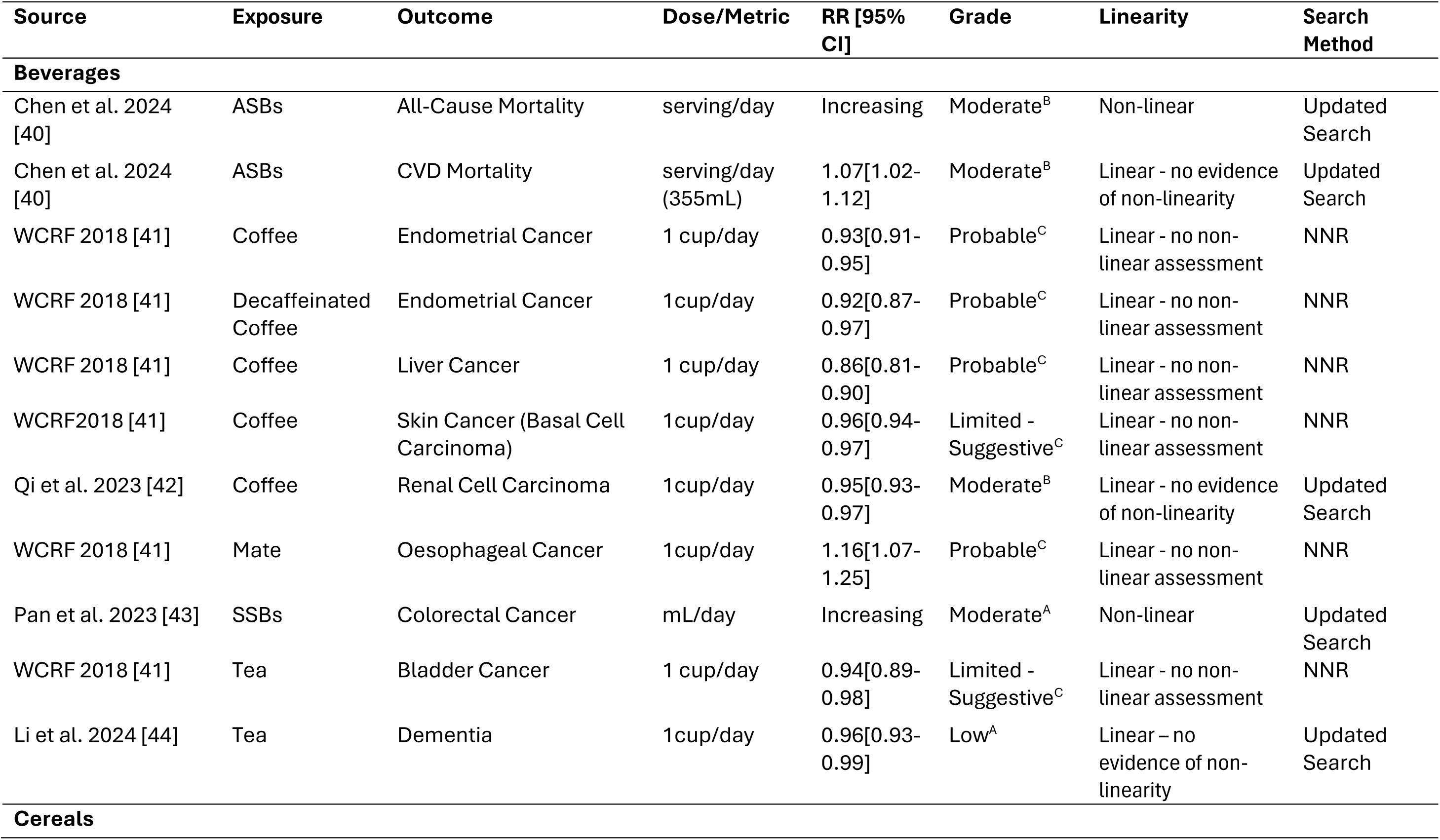
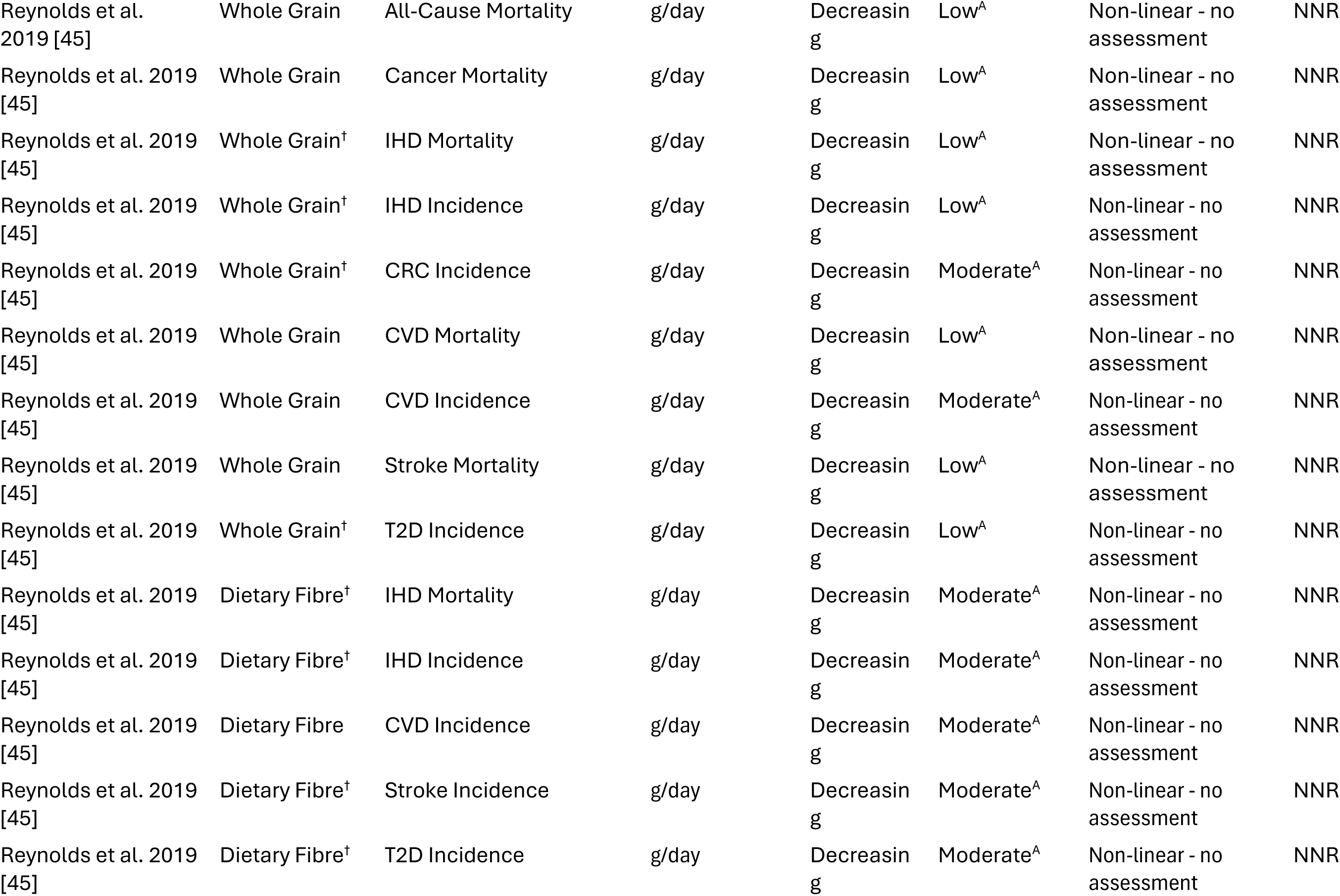
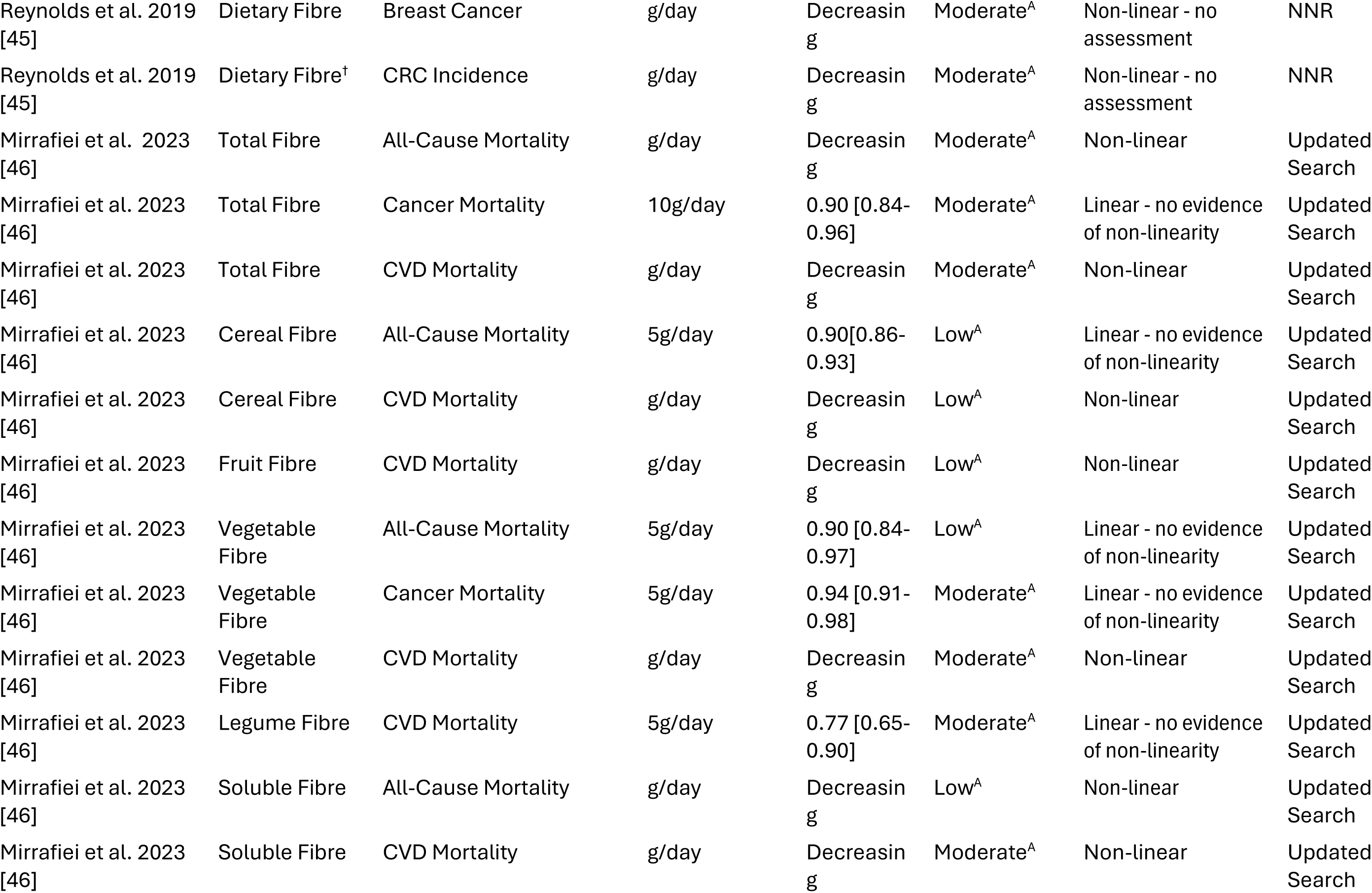
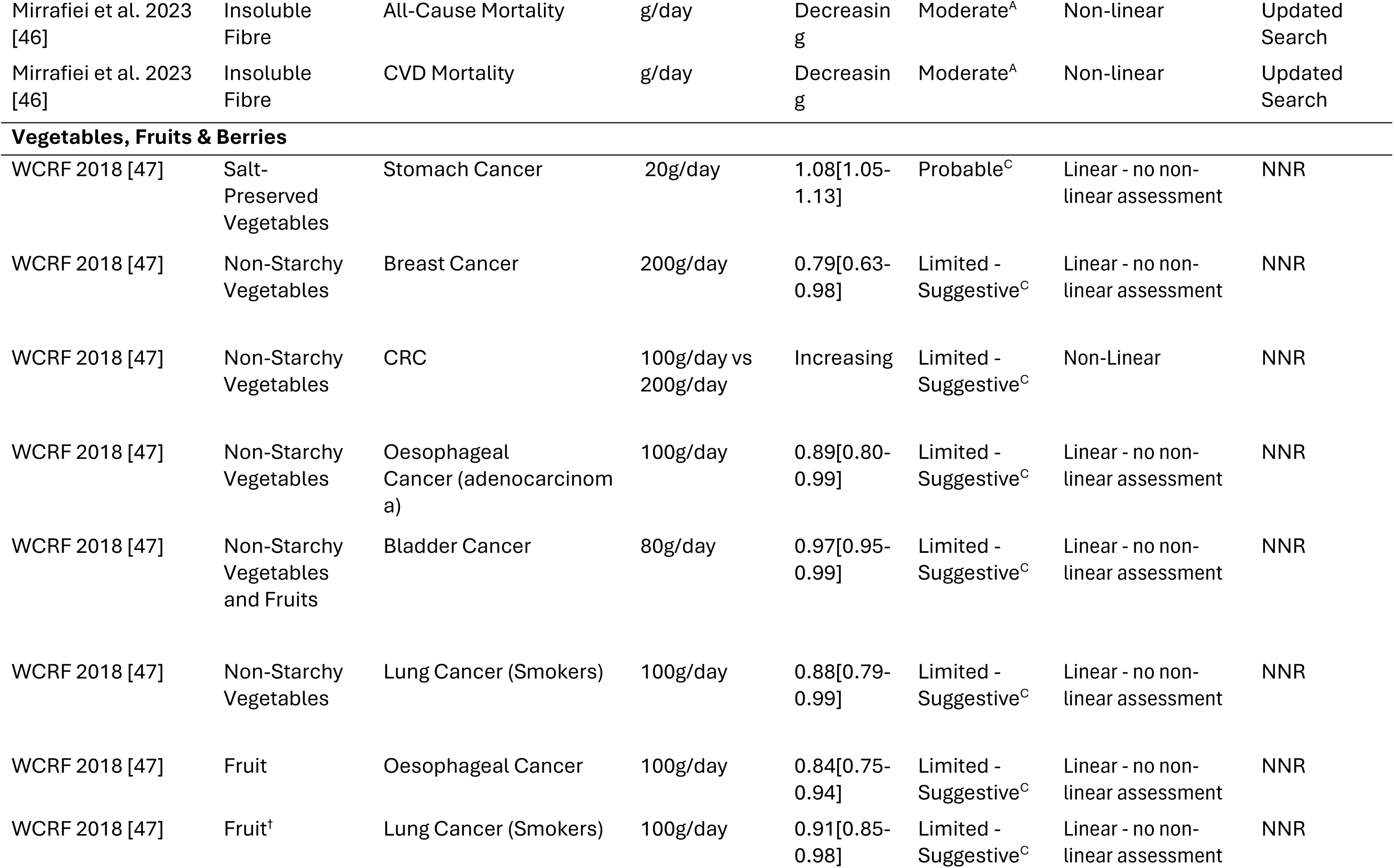
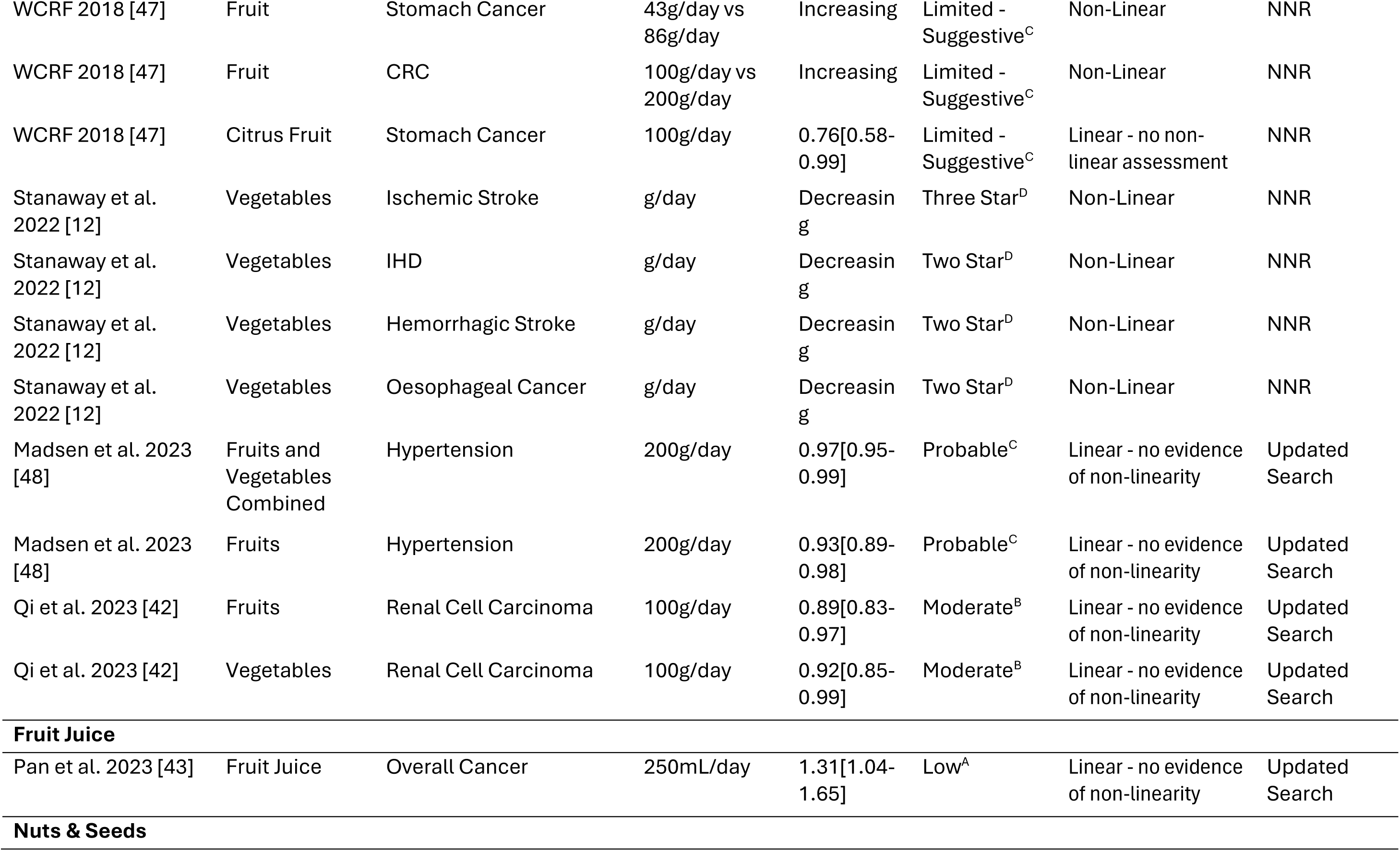
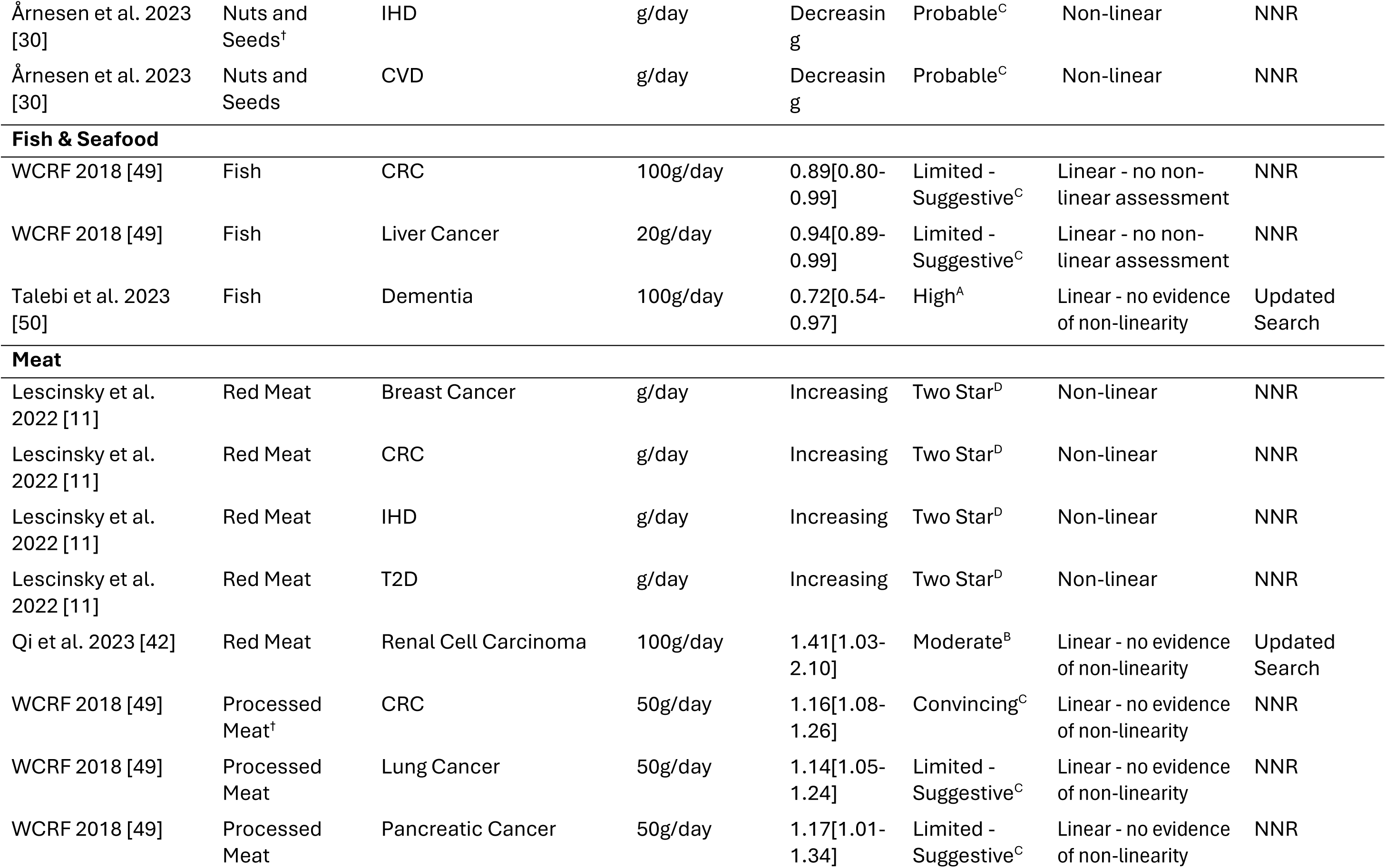
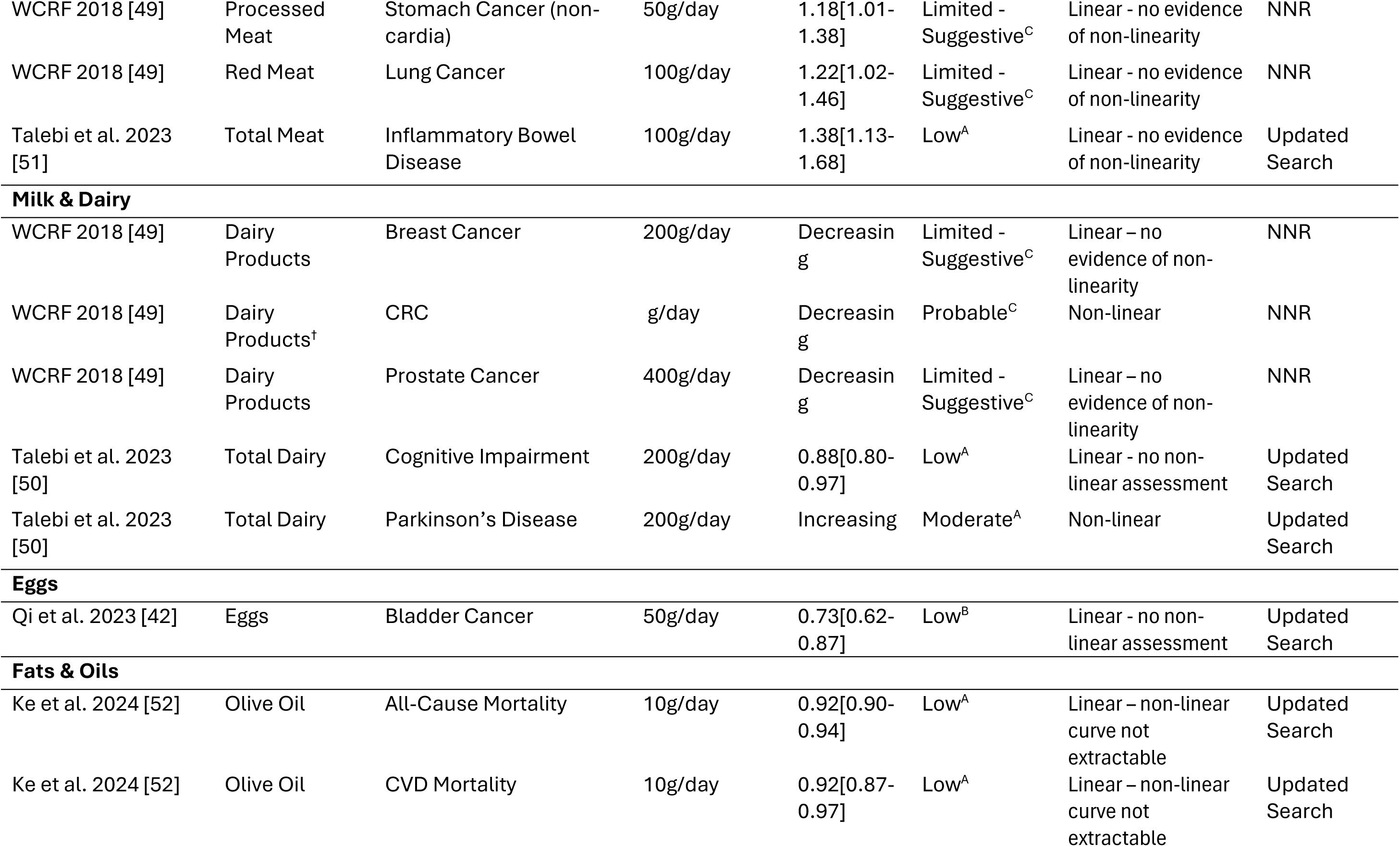
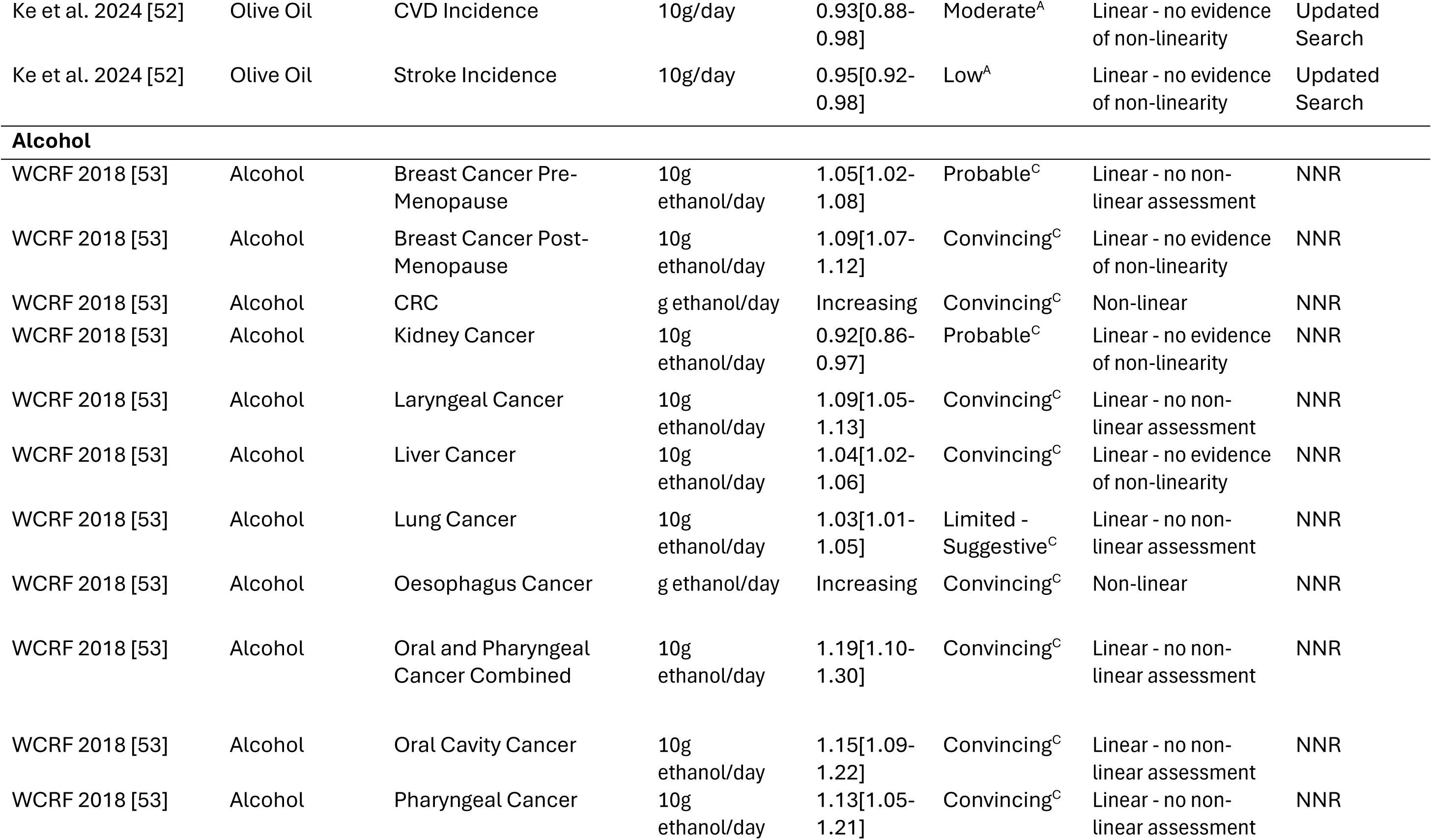
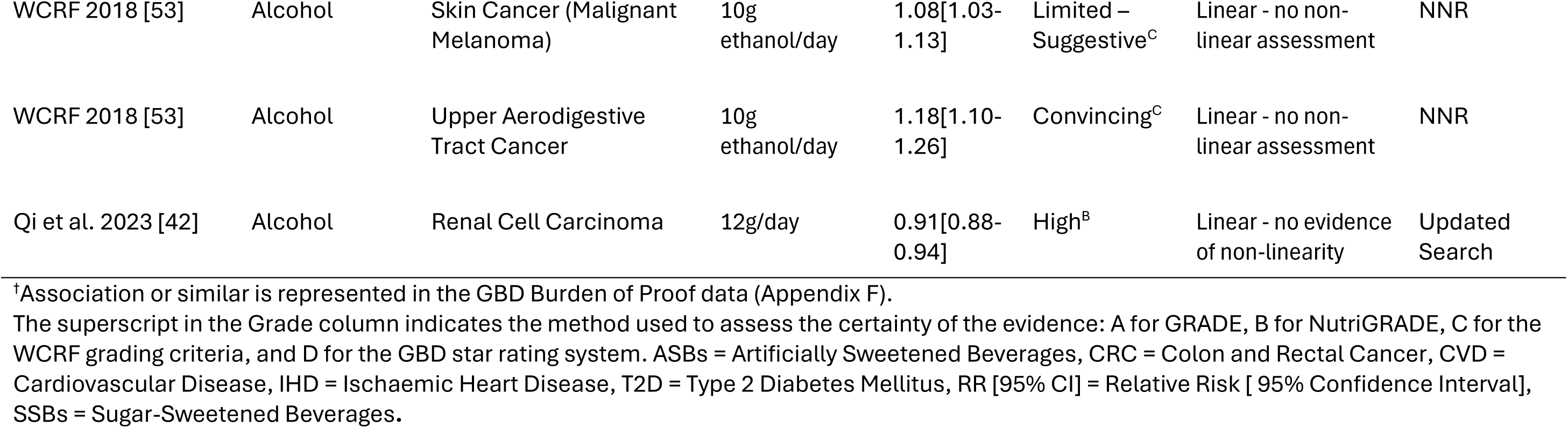
Summary of selected evidence summarised from both NNR2023 and search results.

| Source | Exposure | Outcome | Dose/Metric | RR [95% CI] | Grade | Linearity | Search Method |
| --- | --- | --- | --- | --- | --- | --- | --- |
| **Beverages** |  |  |  |  |  |  |  |
| Chen et al. 2024 [40] | ASBs | All-Cause Mortality | serving/day | Increasing | Moderate<sup>B</sup> | Non-linear | Updated Search |
| Chen et al. 2024 [40] | ASBs | CVD Mortality | serving/day (355mL) | 1.07[1.02-1.12] | Moderate<sup>B</sup> | Linear - no evidence of non-linearity | Updated Search |
| WCRF 2018 [41] | Coffee | Endometrial Cancer | 1 cup/day | 0.93[0.91-0.95] | Probable<sup>C</sup> | Linear - no non-linear assessment | NNR |
| WCRF 2018 [41] | Decaffeinated Coffee | Endometrial Cancer | 1cup/day | 0.92[0.87-0.97] | Probable<sup>C</sup> | Linear - no non-linear assessment | NNR |
| WCRF 2018 [41] | Coffee | Liver Cancer | 1 cup/day | 0.86[0.81-0.90] | Probable<sup>C</sup> | Linear - no non-linear assessment | NNR |
| WCRF2018 [41] | Coffee | Skin Cancer (Basal Cell Carcinoma) | 1cup/day | 0.96[0.94-0.97] | Limited - Suggestive<sup>C</sup> | Linear - no non-linear assessment | NNR |
| Qi et al. 2023 [42] | Coffee | Renal Cell Carcinoma | 1cup/day | 0.95[0.93-0.97] | Moderate<sup>B</sup> | Linear - no evidence of non-linearity | Updated Search |
| WCRF 2018 [41] | Mate | Oesophageal Cancer | 1cup/day | 1.16[1.07-1.25] | Probable<sup>C</sup> | Linear - no non-linear assessment | NNR |
| Pan et al. 2023 [43] | SSBs | Colorectal Cancer | mL/day | Increasing | Moderate<sup>A</sup> | Non-linear | Updated Search |
| WCRF 2018 [41] | Tea | Bladder Cancer | 1 cup/day | 0.94[0.89-0.98] | Limited - Suggestive<sup>C</sup> | Linear - no non-linear assessment | NNR |
| Li et al. 2024 [44] | Tea | Dementia | 1cup/day | 0.96[0.93-0.99] | Low<sup>A</sup> | Linear – no evidence of non-linearity | Updated Search |
| **Cereals** |  |  |  |  |  |  |  |
| Reynolds et al. 2019 [45] | Whole Grain | All-Cause Mortality | g/day | Decreasin<br>g | Low<sup>A</sup> | Non-linear - no<br>assessment | NNR |
| Reynolds et al. 2019 [45] | Whole Grain | Cancer Mortality | g/day | Decreasin<br>g | Low<sup>A</sup> | Non-linear - no<br>assessment | NNR |
| Reynolds et al. 2019 [45] | Whole Grain<sup>†</sup> | IHD Mortality | g/day | Decreasin<br>g | Low<sup>A</sup> | Non-linear - no<br>assessment | NNR |
| Reynolds et al. 2019 [45] | Whole Grain<sup>†</sup> | IHD Incidence | g/day | Decreasin<br>g | Low<sup>A</sup> | Non-linear - no<br>assessment | NNR |
| Reynolds et al. 2019 [45] | Whole Grain<sup>†</sup> | CRC Incidence | g/day | Decreasin<br>g | Moderate<sup>A</sup> | Non-linear - no<br>assessment | NNR |
| Reynolds et al. 2019 [45] | Whole Grain | CVD Mortality | g/day | Decreasin<br>g | Low<sup>A</sup> | Non-linear - no<br>assessment | NNR |
| Reynolds et al. 2019 [45] | Whole Grain | CVD Incidence | g/day | Decreasin<br>g | Moderate<sup>A</sup> | Non-linear - no<br>assessment | NNR |
| Reynolds et al. 2019 [45] | Whole Grain | Stroke Mortality | g/day | Decreasin<br>g | Low<sup>A</sup> | Non-linear - no<br>assessment | NNR |
| Reynolds et al. 2019 [45] | Whole Grain<sup>†</sup> | T2D Incidence | g/day | Decreasin<br>g | Low<sup>A</sup> | Non-linear - no<br>assessment | NNR |
| Reynolds et al. 2019 [45] | Dietary Fibre<sup>†</sup> | IHD Mortality | g/day | Decreasin<br>g | Moderate<sup>A</sup> | Non-linear - no<br>assessment | NNR |
| Reynolds et al. 2019 [45] | Dietary Fibre<sup>†</sup> | IHD Incidence | g/day | Decreasin<br>g | Moderate<sup>A</sup> | Non-linear - no<br>assessment | NNR |
| Reynolds et al. 2019 [45] | Dietary Fibre | CVD Incidence | g/day | Decreasin<br>g | Moderate<sup>A</sup> | Non-linear - no<br>assessment | NNR |
| Reynolds et al. 2019 [45] | Dietary Fibre<sup>†</sup> | Stroke Incidence | g/day | Decreasin<br>g | Moderate<sup>A</sup> | Non-linear - no<br>assessment | NNR |
| Reynolds et al. 2019 [45] | Dietary Fibre<sup>†</sup> | T2D Incidence | g/day | Decreasin<br>g | Moderate<sup>A</sup> | Non-linear - no<br>assessment | NNR |
| Reynolds et al. 2019 [45] | Dietary Fibre | Breast Cancer | g/day | Decreasin<br>g | Moderate<sup>A</sup> | Non-linear - no<br>assessment | NNR |
| Reynolds et al. 2019 [45] | Dietary Fibre<sup>†</sup> | CRC Incidence | g/day | Decreasin<br>g | Moderate<sup>A</sup> | Non-linear - no<br>assessment | NNR |
| Mirrafiei et al. 2023 [46] | Total Fibre | All-Cause Mortality | g/day | Decreasin<br>g | Moderate<sup>A</sup> | Non-linear | Updated<br>Search |
| Mirrafiei et al. 2023 [46] | Total Fibre | Cancer Mortality | 10g/day | 0.90 [0.84-0.96] | Moderate<sup>A</sup> | Linear - no evidence<br>of non-linearity | Updated<br>Search |
| Mirrafiei et al. 2023 [46] | Total Fibre | CVD Mortality | g/day | Decreasin<br>g | Moderate<sup>A</sup> | Non-linear | Updated<br>Search |
| Mirrafiei et al. 2023 [46] | Cereal Fibre | All-Cause Mortality | 5g/day | 0.90[0.86-0.93] | Low<sup>A</sup> | Linear - no evidence<br>of non-linearity | Updated<br>Search |
| Mirrafiei et al. 2023 [46] | Cereal Fibre | CVD Mortality | g/day | Decreasin<br>g | Low<sup>A</sup> | Non-linear | Updated<br>Search |
| Mirrafiei et al. 2023 [46] | Fruit Fibre | CVD Mortality | g/day | Decreasin<br>g | Low<sup>A</sup> | Non-linear | Updated<br>Search |
| Mirrafiei et al. 2023 [46] | Vegetable<br>Fibre | All-Cause Mortality | 5g/day | 0.90 [0.84-0.97] | Low<sup>A</sup> | Linear - no evidence<br>of non-linearity | Updated<br>Search |
| Mirrafiei et al. 2023 [46] | Vegetable<br>Fibre | Cancer Mortality | 5g/day | 0.94 [0.91-0.98] | Moderate<sup>A</sup> | Linear - no evidence<br>of non-linearity | Updated<br>Search |
| Mirrafiei et al. 2023 [46] | Vegetable<br>Fibre | CVD Mortality | g/day | Decreasin<br>g | Moderate<sup>A</sup> | Non-linear | Updated<br>Search |
| Mirrafiei et al. 2023 [46] | Legume Fibre | CVD Mortality | 5g/day | 0.77 [0.65-0.90] | Moderate<sup>A</sup> | Linear - no evidence<br>of non-linearity | Updated<br>Search |
| Mirrafiei et al. 2023 [46] | Soluble Fibre | All-Cause Mortality | g/day | Decreasin<br>g | Low<sup>A</sup> | Non-linear | Updated<br>Search |
| Mirrafiei et al. 2023 [46] | Soluble Fibre | CVD Mortality | g/day | Decreasin<br>g | Moderate<sup>A</sup> | Non-linear | Updated<br>Search |
| Mirrafiei et al. 2023 [46] | Insoluble Fibre | All-Cause Mortality | g/day | Decreasin<br>g | Moderate<sup>A</sup> | Non-linear | Updated Search |
| Mirrafiei et al. 2023 [46] | Insoluble Fibre | CVD Mortality | g/day | Decreasin<br>g | Moderate<sup>A</sup> | Non-linear | Updated Search |
| **Vegetables, Fruits & Berries** |  |  |  |  |  |  |  |
| WCRF 2018 [47] | Salt-Preserved Vegetables | Stomach Cancer | 20g/day | 1.08[1.05-1.13] | Probable<sup>C</sup> | Linear - no non-linear assessment | NNR |
| WCRF 2018 [47] | Non-Starchy Vegetables | Breast Cancer | 200g/day | 0.79[0.63-0.98] | Limited - Suggestive<sup>C</sup> | Linear - no non-linear assessment | NNR |
| WCRF 2018 [47] | Non-Starchy Vegetables | CRC | 100g/day vs 200g/day | Increasing | Limited - Suggestive<sup>C</sup> | Non-Linear | NNR |
| WCRF 2018 [47] | Non-Starchy Vegetables | Oesophageal Cancer (adenocarcinoma) | 100g/day | 0.89[0.80-0.99] | Limited - Suggestive<sup>C</sup> | Linear - no non-linear assessment | NNR |
| WCRF 2018 [47] | Non-Starchy Vegetables and Fruits | Bladder Cancer | 80g/day | 0.97[0.95-0.99] | Limited - Suggestive<sup>C</sup> | Linear - no non-linear assessment | NNR |
| WCRF 2018 [47] | Non-Starchy Vegetables | Lung Cancer (Smokers) | 100g/day | 0.88[0.79-0.99] | Limited - Suggestive<sup>C</sup> | Linear - no non-linear assessment | NNR |
| WCRF 2018 [47] | Fruit | Oesophageal Cancer | 100g/day | 0.84[0.75-0.94] | Limited - Suggestive<sup>C</sup> | Linear - no non-linear assessment | NNR |
| WCRF 2018 [47] | Fruit<sup>†</sup> | Lung Cancer (Smokers) | 100g/day | 0.91[0.85-0.98] | Limited - Suggestive<sup>C</sup> | Linear - no non-linear assessment | NNR |
| WCRF 2018 [47] | Fruit | Stomach Cancer | 43g/day vs 86g/day | Increasing | Limited - Suggestive<sup>C</sup> | Non-Linear | NNR |
| WCRF 2018 [47] | Fruit | CRC | 100g/day vs 200g/day | Increasing | Limited - Suggestive<sup>C</sup> | Non-Linear | NNR |
| WCRF 2018 [47] | Citrus Fruit | Stomach Cancer | 100g/day | 0.76[0.58-0.99] | Limited - Suggestive<sup>C</sup> | Linear - no non-linear assessment | NNR |
| Stanaway et al. 2022 [12] | Vegetables | Ischemic Stroke | g/day | Decreasing | Three Star<sup>D</sup> | Non-Linear | NNR |
| Stanaway et al. 2022 [12] | Vegetables | IHD | g/day | Decreasing | Two Star<sup>D</sup> | Non-Linear | NNR |
| Stanaway et al. 2022 [12] | Vegetables | Hemorrhagic Stroke | g/day | Decreasing | Two Star<sup>D</sup> | Non-Linear | NNR |
| Stanaway et al. 2022 [12] | Vegetables | Oesophageal Cancer | g/day | Decreasing | Two Star<sup>D</sup> | Non-Linear | NNR |
| Madsen et al. 2023 [48] | Fruits and Vegetables Combined | Hypertension | 200g/day | 0.97[0.95-0.99] | Probable<sup>C</sup> | Linear - no evidence of non-linearity | Updated Search |
| Madsen et al. 2023 [48] | Fruits | Hypertension | 200g/day | 0.93[0.89-0.98] | Probable<sup>C</sup> | Linear - no evidence of non-linearity | Updated Search |
| Qi et al. 2023 [42] | Fruits | Renal Cell Carcinoma | 100g/day | 0.89[0.83-0.97] | Moderate<sup>B</sup> | Linear - no evidence of non-linearity | Updated Search |
| Qi et al. 2023 [42] | Vegetables | Renal Cell Carcinoma | 100g/day | 0.92[0.85-0.99] | Moderate<sup>B</sup> | Linear - no evidence of non-linearity | Updated Search |
| **Fruit Juice** |  |  |  |  |  |  |  |
| Pan et al. 2023 [43] | Fruit Juice | Overall Cancer | 250mL/day | 1.31[1.04-1.65] | Low<sup>A</sup> | Linear - no evidence of non-linearity | Updated Search |
| **Nuts & Seeds** |  |  |  |  |  |  |  |
| Årnesen et al. 2023 [30] | Nuts and Seeds<sup>†</sup> | IHD | g/day | Decreasin<br>g | Probable<sup>C</sup> | Non-linear | NNR |
| Årnesen et al. 2023 [30] | Nuts and Seeds | CVD | g/day | Decreasin<br>g | Probable<sup>C</sup> | Non-linear | NNR |
| **Fish & Seafood** |  |  |  |  |  |  |  |
| WCRF 2018 [49] | Fish | CRC | 100g/day | 0.89[0.80-0.99] | Limited - Suggestive<sup>C</sup> | Linear - no non-linear assessment | NNR |
| WCRF 2018 [49] | Fish | Liver Cancer | 20g/day | 0.94[0.89-0.99] | Limited - Suggestive<sup>C</sup> | Linear - no non-linear assessment | NNR |
| Talebi et al. 2023 [50] | Fish | Dementia | 100g/day | 0.72[0.54-0.97] | High<sup>A</sup> | Linear - no evidence of non-linearity | Updated Search |
| **Meat** |  |  |  |  |  |  |  |
| Lescinsky et al. 2022 [11] | Red Meat | Breast Cancer | g/day | Increasing | Two Star<sup>D</sup> | Non-linear | NNR |
| Lescinsky et al. 2022 [11] | Red Meat | CRC | g/day | Increasing | Two Star<sup>D</sup> | Non-linear | NNR |
| Lescinsky et al. 2022 [11] | Red Meat | IHD | g/day | Increasing | Two Star<sup>D</sup> | Non-linear | NNR |
| Lescinsky et al. 2022 [11] | Red Meat | T2D | g/day | Increasing | Two Star<sup>D</sup> | Non-linear | NNR |
| Qi et al. 2023 [42] | Red Meat | Renal Cell Carcinoma | 100g/day | 1.41[1.03-2.10] | Moderate<sup>B</sup> | Linear - no evidence of non-linearity | Updated Search |
| WCRF 2018 [49] | Processed Meat<sup>†</sup> | CRC | 50g/day | 1.16[1.08-1.26] | Convincing<sup>C</sup> | Linear - no evidence of non-linearity | NNR |
| WCRF 2018 [49] | Processed Meat | Lung Cancer | 50g/day | 1.14[1.05-1.24] | Limited - Suggestive<sup>C</sup> | Linear - no evidence of non-linearity | NNR |
| WCRF 2018 [49] | Processed Meat | Pancreatic Cancer | 50g/day | 1.17[1.01-1.34] | Limited - Suggestive<sup>C</sup> | Linear - no evidence of non-linearity | NNR |
| WCRF 2018 [49] | Processed Meat | Stomach Cancer (non-cardia) | 50g/day | 1.18[1.01-1.38] | Limited - Suggestive<sup>C</sup> | Linear - no evidence of non-linearity | NNR |
| WCRF 2018 [49] | Red Meat | Lung Cancer | 100g/day | 1.22[1.02-1.46] | Limited - Suggestive<sup>C</sup> | Linear - no evidence of non-linearity | NNR |
| Talebi et al. 2023 [51] | Total Meat | Inflammatory Bowel Disease | 100g/day | 1.38[1.13-1.68] | Low<sup>A</sup> | Linear - no evidence of non-linearity | Updated Search |
| **Milk & Dairy** |  |  |  |  |  |  |  |
| WCRF 2018 [49] | Dairy Products | Breast Cancer | 200g/day | Decreasing | Limited - Suggestive<sup>C</sup> | Linear – no evidence of non-linearity | NNR |
| WCRF 2018 [49] | Dairy Products<sup>†</sup> | CRC | g/day | Decreasing | Probable<sup>C</sup> | Non-linear | NNR |
| WCRF 2018 [49] | Dairy Products | Prostate Cancer | 400g/day | Decreasing | Limited - Suggestive<sup>C</sup> | Linear – no evidence of non-linearity | NNR |
| Talebi et al. 2023 [50] | Total Dairy | Cognitive Impairment | 200g/day | 0.88[0.80-0.97] | Low<sup>A</sup> | Linear - no non-linear assessment | Updated Search |
| Talebi et al. 2023 [50] | Total Dairy | Parkinson's Disease | 200g/day | Increasing | Moderate<sup>A</sup> | Non-linear | Updated Search |
| **Eggs** |  |  |  |  |  |  |  |
| Qi et al. 2023 [42] | Eggs | Bladder Cancer | 50g/day | 0.73[0.62-0.87] | Low<sup>B</sup> | Linear - no non-linear assessment | Updated Search |
| **Fats & Oils** |  |  |  |  |  |  |  |
| Ke et al. 2024 [52] | Olive Oil | All-Cause Mortality | 10g/day | 0.92[0.90-0.94] | Low<sup>A</sup> | Linear – non-linear curve not extractable | Updated Search |
| Ke et al. 2024 [52] | Olive Oil | CVD Mortality | 10g/day | 0.92[0.87-0.97] | Low<sup>A</sup> | Linear – non-linear curve not extractable | Updated Search |
| Ke et al. 2024 [52] | Olive Oil | CVD Incidence | 10g/day | 0.93[0.88-0.98] | Moderate<sup>A</sup> | Linear - no evidence of non-linearity | Updated Search |
| Ke et al. 2024 [52] | Olive Oil | Stroke Incidence | 10g/day | 0.95[0.92-0.98] | Low<sup>A</sup> | Linear - no evidence of non-linearity | Updated Search |
| Alcohol |  |  |  |  |  |  |  |
| WCRF 2018 [53] | Alcohol | Breast Cancer Pre-Menopause | 10g ethanol/day | 1.05[1.02-1.08] | Probable<sup>C</sup> | Linear - no non-linear assessment | NNR |
| WCRF 2018 [53] | Alcohol | Breast Cancer Post-Menopause | 10g ethanol/day | 1.09[1.07-1.12] | Convincing<sup>C</sup> | Linear - no evidence of non-linearity | NNR |
| WCRF 2018 [53] | Alcohol | CRC | g ethanol/day | Increasing | Convincing<sup>C</sup> | Non-linear | NNR |
| WCRF 2018 [53] | Alcohol | Kidney Cancer | 10g ethanol/day | 0.92[0.86-0.97] | Probable<sup>C</sup> | Linear - no evidence of non-linearity | NNR |
| WCRF 2018 [53] | Alcohol | Laryngeal Cancer | 10g ethanol/day | 1.09[1.05-1.13] | Convincing<sup>C</sup> | Linear - no non-linear assessment | NNR |
| WCRF 2018 [53] | Alcohol | Liver Cancer | 10g ethanol/day | 1.04[1.02-1.06] | Convincing<sup>C</sup> | Linear - no evidence of non-linearity | NNR |
| WCRF 2018 [53] | Alcohol | Lung Cancer | 10g ethanol/day | 1.03[1.01-1.05] | Limited - Suggestive<sup>C</sup> | Linear - no non-linear assessment | NNR |
| WCRF 2018 [53] | Alcohol | Oesophagus Cancer | g ethanol/day | Increasing | Convincing<sup>C</sup> | Non-linear | NNR |
| WCRF 2018 [53] | Alcohol | Oral and Pharyngeal Cancer Combined | 10g ethanol/day | 1.19[1.10-1.30] | Convincing<sup>C</sup> | Linear - no non-linear assessment | NNR |
| WCRF 2018 [53] | Alcohol | Oral Cavity Cancer | 10g ethanol/day | 1.15[1.09-1.22] | Convincing<sup>C</sup> | Linear - no non-linear assessment | NNR |
| WCRF 2018 [53] | Alcohol | Pharyngeal Cancer | 10g ethanol/day | 1.13[1.05-1.21] | Convincing<sup>C</sup> | Linear - no non-linear assessment | NNR |
| WCRF 2018 [53] | Alcohol | Skin Cancer (Malignant Melanoma) | 10g ethanol/day | 1.08[1.03-1.13] | Limited – Suggestive<sup>C</sup> | Linear - no non-linear assessment | NNR |
| WCRF 2018 [53] | Alcohol | Upper Aerodigestive Tract Cancer | 10g ethanol/day | 1.18[1.10-1.26] | Convincing<sup>C</sup> | Linear - no non-linear assessment | NNR |
| Qi et al. 2023 [42] | Alcohol | Renal Cell Carcinoma | 12g/day | 0.91[0.88-0.94] | High<sup>B</sup> | Linear - no evidence of non-linearity | Updated Search |

<sup>†</sup>Association or similar is represented in the GBD Burden of Proof data (Appendix F).
The superscript in the Grade column indicates the method used to assess the certainty of the evidence: A for GRADE, B for NutriGRADE, C for the WCRF grading criteria, and D for the GBD star rating system. ASBs = Artificially Sweetened Beverages, CRC = Colon and Rectal Cancer, CVD = Cardiovascular Disease, IHD = Ischaemic Heart Disease, T2D = Type 2 Diabetes Mellitus, RR [95% CI] = Relative Risk [ 95% Confidence Interval], SSBs = Sugar-Sweetened Beverages.

From the original NNR2023 qualified systematic reviews for food-based dietary guidelines (n=27), 11 were screened for individual risk-outcome pairs [5]. From these studies, 70 risk-outcome associations were identified (Figure 1). The updated search resulted in 15 included studies (Figure 1), and from which 36 risk-outcome pairs were included (coming from 9 of the 15 included studies). Five of the selected pairs were conflicting in the sense that they had the same outcome and exposure but came from different studies. Following the critical comparison, a single pair was selected for each exposure-outcome (Appendix D, Table D1 – D5). This left a total of 101 risk-outcome associations included in the database (Figure 1).

**Figure 1.**
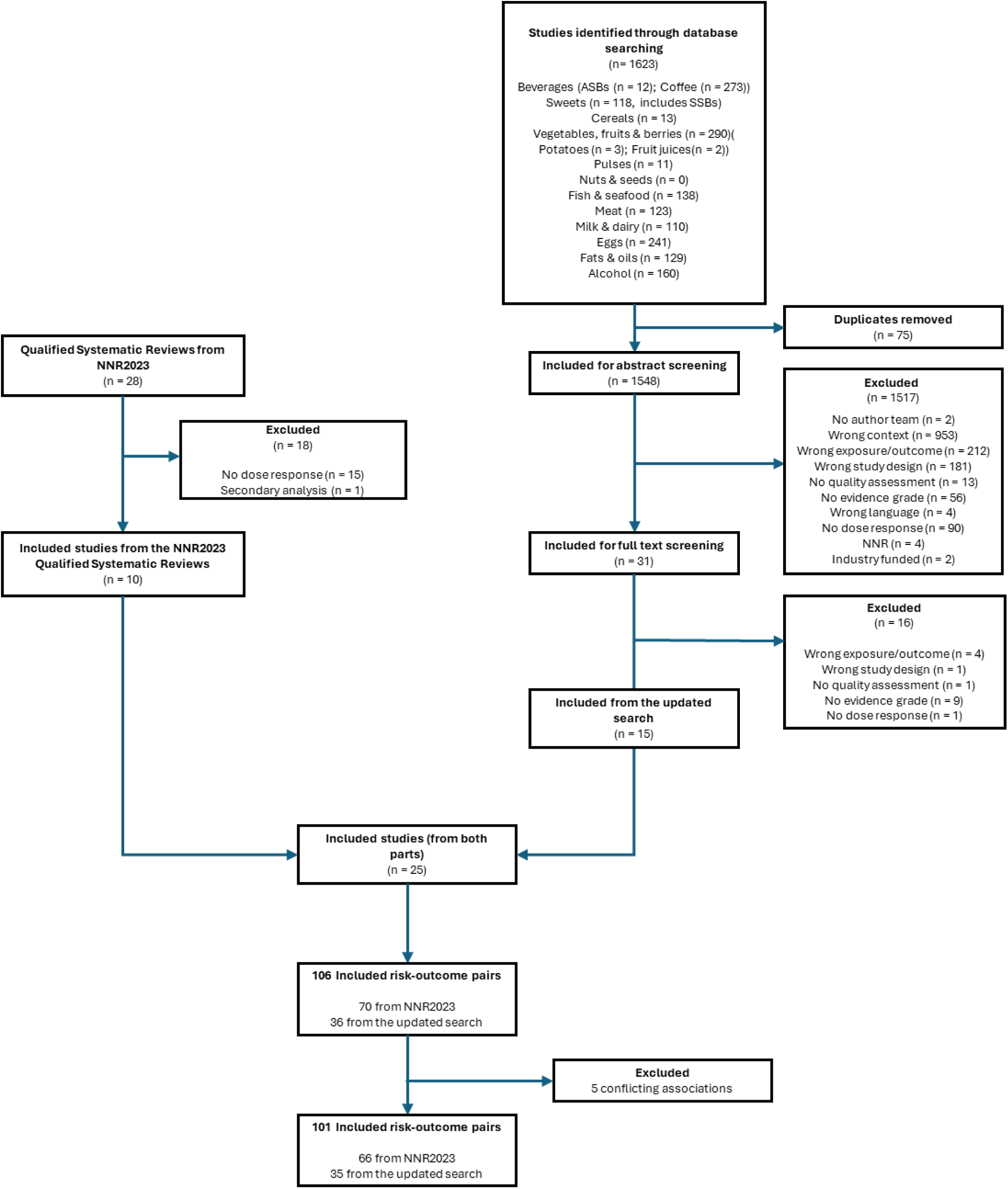
Flow diagram of the article screening and selection process. ASBs = Artificially Sweetened Beverages, NNR = Nordic Nutrition Recommendations, SSBs = Sugar and Sweetened Beverages.

Figure 2 depicts the qualitative mapping conducted to compare the descriptive features of the different grading systems and to determine a logical cut-off point across the different approaches. In total, 52 of the 101 included risk-outcome pairs had an evidence grade at or above moderate (GRADE and NutriGRADE), probable (WCRF), or three stars (GBD Burden of Proof). Out of the 44 dietary risk-outcome pairs in the GBD 2021, 20 pairs were rated two or more stars and included food-based exposures (nutrient-based dietary risk factors were excluded), and from these three were rated three stars (Appendix F, Table F1).

**Figure 2:**
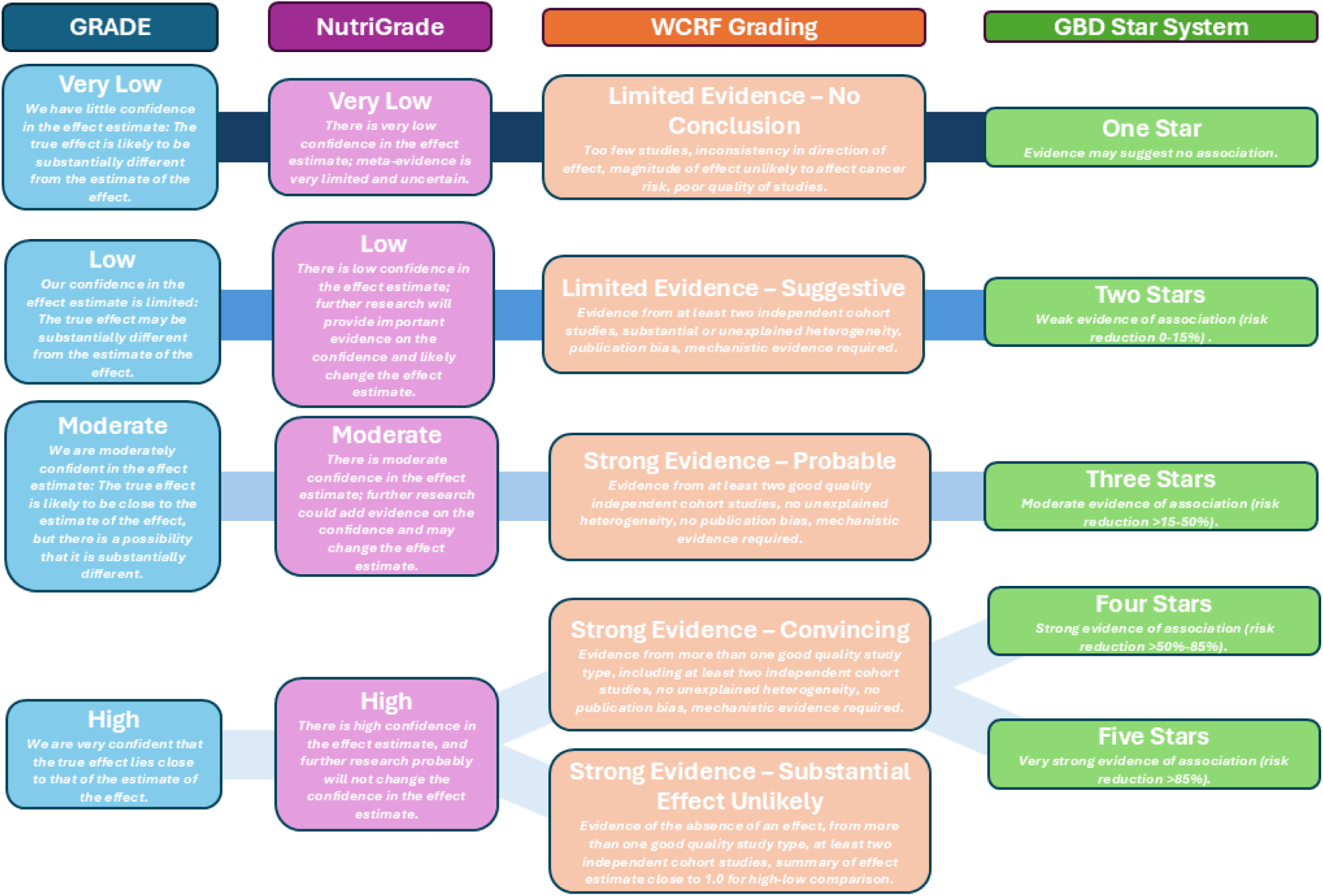
Qualitative mapping of grading systems GRADE [54], NutriGrade [33], WCRF Grading [34], and GBD Star System [55]. Equivalent levels are drawn based on qualitative descriptions and relative position within the grading system. There are known misalignments here (see Discussion of main paper).

Non-linear curves were extracted from visual graphs, fit with piecewise functions, and were compiled alongside the linear associations in the database [39]. An example of the curve extraction and function fitting process can be found in Figure 3.

**Figure 3.**
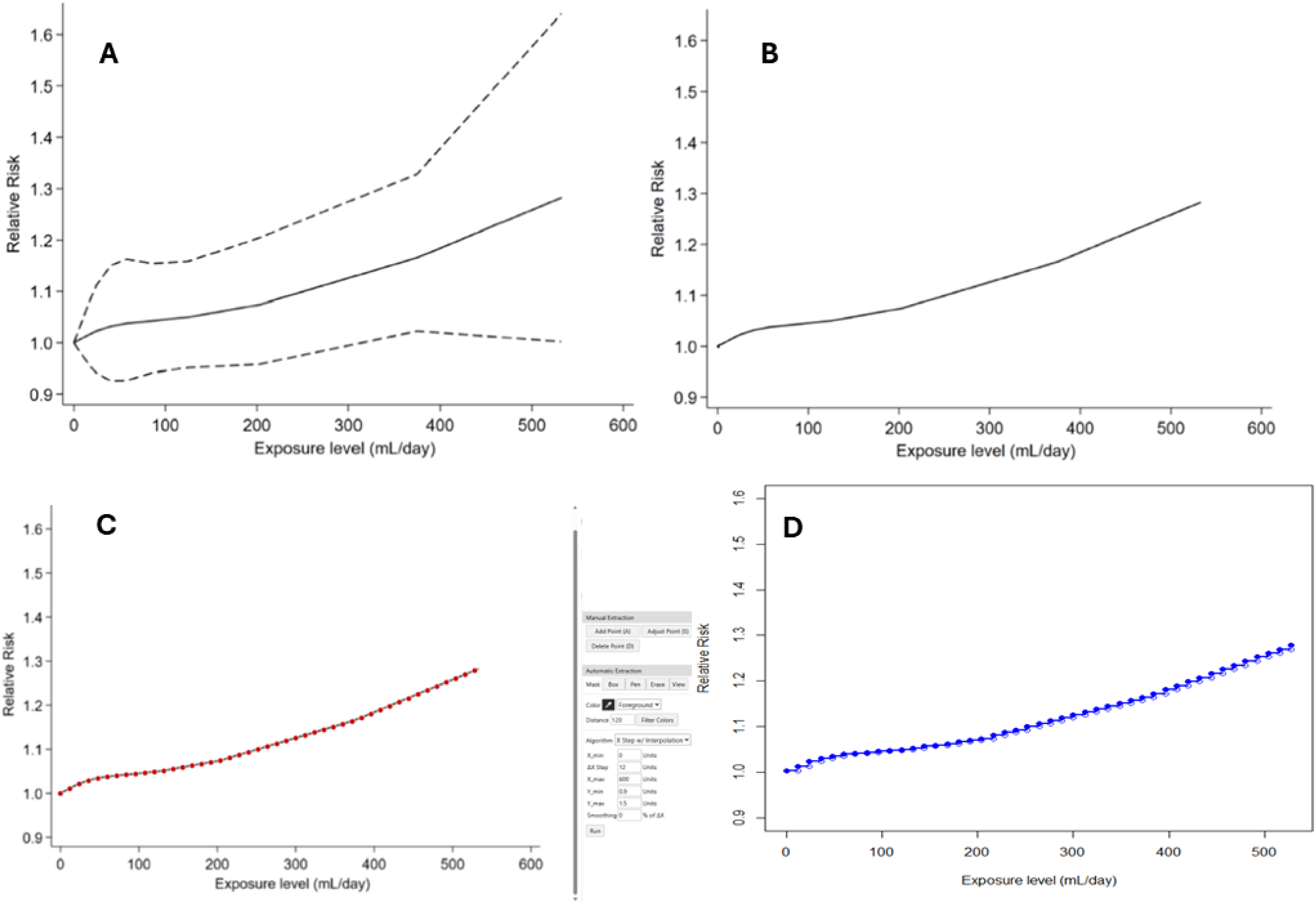
Example non-linear dose-response curve extraction of the association between SSBs and CRC from Pan et al. 2023 [43]. A) The original curve, reproduced with permission. B) The curve image stripped of additional information. C) Point extraction from WebPlotDigitiser [37]. D) Fitted piecewise constant function to estimate the curve.

## Discussion

We selected and compiled a total of 101 dietary risk-outcome pairs in our database, of which 35 came from the updated search. Extraction of dose-response curves for the selected risk-outcome pairs, particularly non-linear associations (n=44), was completed using a novel and efficient method that has been made openly accessible for future applications. The process of updating, selecting, and extracting dietary risk-outcome pairs and their associated dose-response curves from existing epidemiological data highlighted and confirmed many of the known challenges modellers and researchers face in applying these data to health impact assessments. Recommendations for overcoming some of these challenges are discussed throughout the following section and are summarised in Table 3.

**Table 3.**
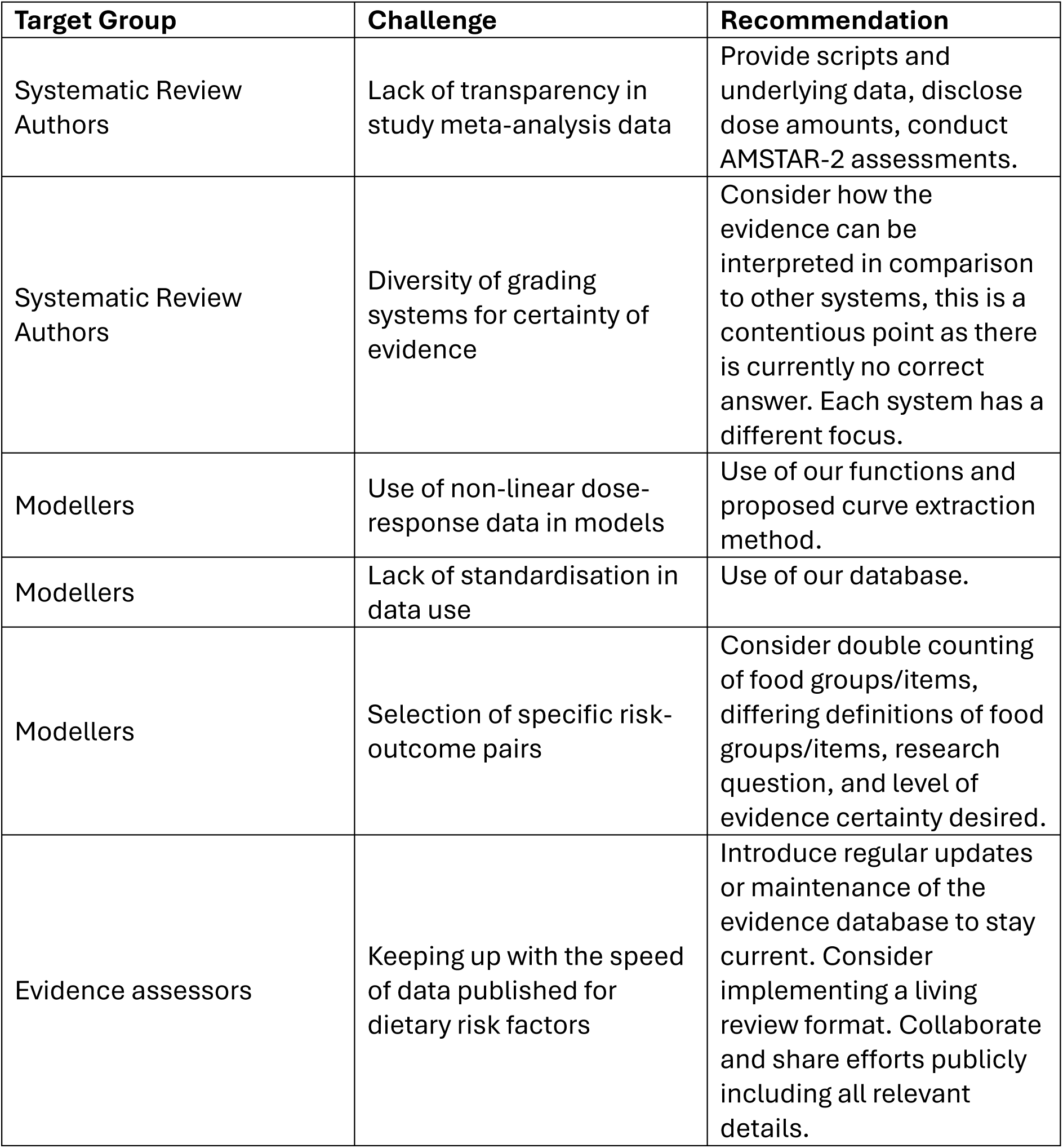
Recommendations to stakeholders for improved health impact assessments.

| Target Group | Challenge | Recommendation |
| --- | --- | --- |
| Systematic Review Authors | Lack of transparency in study meta-analysis data | Provide scripts and underlying data, disclose dose amounts, conduct AMSTAR-2 assessments. |
| Systematic Review Authors | Diversity of grading systems for certainty of evidence | Consider how the evidence can be interpreted in comparison to other systems, this is a contentious point as there is currently no correct answer. Each system has a different focus. |
| Modellers | Use of non-linear dose-response data in models | Use of our functions and proposed curve extraction method. |
| Modellers | Lack of standardisation in data use | Use of our database. |
| Modellers | Selection of specific risk-outcome pairs | Consider double counting of food groups/items, differing definitions of food groups/items, research question, and level of evidence certainty desired. |
| Evidence assessors | Keeping up with the speed of data published for dietary risk factors | Introduce regular updates or maintenance of the evidence database to stay current. Consider implementing a living review format. Collaborate and share efforts publicly including all relevant details. |

### Comparisons of Data Sources

Comparing our database to other collections of risk-outcome pairs, only moderate alignment between dietary risk-outcome pairs in our database and the GBD study was observed. For example, the associations between type 2 diabetes and ischaemic heart disease and processed meat which exist in the GBD data were not found in the NNR2023 and update data. Only around half of the 23 included GBD risk-outcome pairs were represented in the NNR2023 data (excluding GBD pairs already included in NNR2023) and the certainty of the evidence varied [6,11,12]. Considering whole grain intake as the exposure, the risk of ischaemic heart disease was found in both data sources [6,45]; however, the strength of the evidence in GBD was rated as three stars while it received a “low” on the GRADE scale in the systematic review by Reynolds et al. [45,56]. Conversely, numerous associations were included in our search that were not present in the GBD data such as dairy intake and Parkinson’s disease, red meat intake and renal cell carcinoma, and all of the associations with alcohol [41,42,53]. It was also difficult to compare GBD dietary risk factors with other sources of data due to the level estimates were made at (i.e. food group for GBD but food item level in NNR in some instances). In the GBD, seafood omega-3 is a dietary risk factor, but in other sources, determining if omega-3 came from only seafood or if food items of fish or seafood were comparable was difficult to determine [5,56]. These discrepancies give rise to significant questions regarding the selection process employed to include studies in the two approaches and the lack of standardisation in the field.

Our present study yielded intriguing findings, with some unexpected contrasts to established literature beyond the GBD. If considering only pairs with certainty ratings of at least “probable”; “moderate” or three-star associations, around half of the associations would have been excluded, underscoring the relatively low overall strength of evidence available in the current literature, despite the abundance of systematic reviews and meta-analyses identified. Based on this “higher” threshold, our findings indicate that red meat would only be associated with renal cell carcinoma. This contrasts sharply with existing research qualified as “high” quality of evidence (red and processed meat and type 2 diabetes), or “moderate” (red and processed meat and coronary heart disease, colorectal cancer and all-cause mortality) from prominent meta-analyses such as those from Schwingshackl et al. and Bechthold et al. [57–60]. The same applies to processed meat, which is widely recognised as a risk factor for various diseases [61], but would only have been included for its association with the risk of developing colorectal cancer under a three-star threshold. There were examples of risk-outcome pairs that aligned with previous evidence such as the association between whole grain, vegetables, or red meat and ischaemic heart disease, as also seen in Bechthold et al., but alignment especially in gradings of certainty was rather rare [57].

Moreover, no associations were found between legumes as a whole food and any health outcomes that met the inclusion criteria, despite a widely recognised protective effect against various non-communicable diseases (i.e. in studies lacking evidence grading or dose-response analysis, and in multiple dietary guidelines) [62–64]. This finding is, however, consistent with a recent systematic review and meta-analysis which also failed to identify strong quantitative associations between legumes and type 2 diabetes or cardiovascular diseases [65]. A key challenge noted in the literature is that legume consumption in Western populations is typically very low—often below the thresholds necessary to detect meaningful associations at the population level [66]. Current recommendations to increase legume intake are based on their contribution to nutrients in the diet (e.g. as a substitute for animal-based protein) and protective effects of fibre rather than their direct association of legumes with negative health outcomes [67]. The discrepancies highlight significant differences in the evaluation and classification of dietary risk-outcome associations, raising questions about methodological variations across studies and the implications for dietary health impact assessments.

### Addressing Challenges for Health Impact Assessments

The selection and use of data for the purpose of dietary health impact assessments poses multiple challenges for modellers and researchers. While we have provided all the available data according to our protocol, modellers must still decide on which specific pairs to use from the database based on a variety of factors including modelling requirements and the desired level of certainty of evidence (i.e. a more conservative cut-off point as seen in previous modelling studies) [68–70]. There are also considerations to be made for the lack of assessment for methodology quality (e.g. using AMSTAR-2), avoidance of double-counting food groups or items (e.g. whole grain and dietary fibre) when modelling risk outcomes, and the publication bias that may influence this field. We also faced challenges in determining dose size for certain food items, for example measurement in cups could refer to a US cup (240mL), a UK cup (250mL), a WCRF defined cup (200mL), or another measure entirely, as dose metrics were often not specified in the studies [71].

One of the prominent challenges in evaluating the evidence from various meta-analyses is the diversity of grading systems that exist, which are often difficult to compare. GRADE is a widely used and well-recognised framework for assessing the quality of evidence [72]; however, it has been criticized for classifying all observational studies as low quality, prior to any potential upgrades, which is a challenge in nutritional research where randomised controlled trials are not often feasible [28,73]. This is one reason why the NutriGRADE system was developed to provide a more nuanced approach to grading evidence, particularly for cohort studies [33]. Similarly, WCRF places a significant emphasis on biological plausibility in its grading [34], which is not a typical focus of GRADE or NutriGRADE, further contributing to the complexity of comparing evidence. The GBD uses a different system entirely, combining various factors into the risk outcome score which is then converted into the Burden of Proof Star Rating system [55]. While this approach is easy to understand, especially for the layperson, it is not comparable with other systems, as it uses different scales for positive and negative associations and considers magnitude of association as a main determinant [55]. For our purposes, we attempted to align the grading systems based on their qualitative descriptions, which has not been discussed previously, but in doing so, we acknowledge the limitations knowing the systems are not objectively aligned in their judgements of evidence [33]. Selecting data of the highest quality remains contentious, given the differing assessment criteria for certainty.

Meta-analyses require significant time, resources, and computational commitments, which many researchers — whose purpose is to conduct health impact assessments — do not have the capacity for. In building our database we attempted to overcome many of these known challenges not by being exhaustive, but rather as comprehensive as possible within the constraints. The focus was on ensuring that the data collected is transparent, easily accessible, and replicable, allowing other researchers to reuse and build on the database in a systematic and reproducible way.

### Limitations

Our database exclusively selected associations from meta-analyses with dose-response data, which could introduce selection bias by not considering the contributions of systematic reviews without meta-analyses. This may have omitted evidence not yet documented in a meta-analysis, particularly emerging evidence from smaller or early-phase studies, exemplifying the benefit of frequent updating and regular maintenance of the database to maintain currency.

While the approach of including only commissioned reviews in the qualified systematic reviews sought to ensure methodological rigor and independence, it may have inadvertently excluded high-quality scientific studies that could provide valuable insights [14]. We consider that, in the absence of a conflict of interest and industry funding, non-commissioned systematic reviews can be of significant quality and value in our dietary risk-outcome pair database. As our focus is on constructing a usable database of epidemiological data and not the scientific basis for national dietary guidelines, we omitted this criterion. Since we were starting with the NNR2023 qualified systematic reviews as our baseline, studies that may have been included previously without the commissioning criterion were not represented. There may have therefore been key sources missed, and this bias may partially contribute to the discrepancies between GBD results and our constructed database.

Another limitation is related to the extraction of non-linear dose-response curves. Despite attempts to obtain the curve data directly from the authors of included studies, very few responses were received, and even fewer data. As a result, we developed a novel approach to easily estimate non-linear curves for use in modelling projects. An online graphing tool was used to automatically extract data points from the published curves, which is associated with potential loss of precision and accuracy as the process relies on graphical estimation rather than direct access to raw data. There is also room for human error in calibration of the axis and the assumptions made for data not displayed between zero and the start of points, or after the observable range, which may not accurately describe the relationships. While imperfect, efforts were made to keep the estimation procedure standard and as accurate to the curves’ behaviour as possible, providing reasonable estimation.

### Conclusions

To improve the accuracy and comprehensiveness of dietary risk assessments and manage the overwhelming surge of new evidence in dietary risk factors and modelling, ensuring regular updates of key data sources is essential. Promoting greater alignment between key institutions, such as GBD, WCRF, and NNR, could improve the consistency of dietary risk assessments by integrating a wider range of data sources and methodologies. Our further recommendations for various stakeholders can be found in Table 3. Finally, aligning or quantitative differentiation of grading systems is crucial, as using multiple frameworks—such as the four in this review—can complicate comparisons and the interpretation of evidence.

Selection of dietary risk data poses both practical and theoretical challenges for researchers, especially those responsible for providing decision-makers with rapid and robust evidence. There is no ultimately correct way to select or extract dose-response data for these modelling initiatives. Our development of a dietary risk factor database that can be updated and replicated aimed to overcome some of these challenges and allow for better alignment in the field. It is our intention to now use this aligned data in assessments and share the database with other researchers embarking on similar projects. Improved alignment should allow for improved comparability, strength of results, and robustness of findings in terms of dietary assessment.

## Data Availability

All data produced in the study are available online at Zenodo.

https://doi.org/10.5281/zenodo.16881749

## Funding

This work was partly funded by the Danish Ministry of Food, Agriculture and Fisheries (AEJ). Funders had no role in the design, conduct, writing, or publication of this work.

## Conflicts of Interest

The authors declare no conflicts of interest.

## Notes

### Competing Interest Statement

The authors have declared no competing interest.

### Funding Statement

This study was partly funded by the Danish Ministry of Food, Agriculture and Fisheries (AEJ).

