## Supplementary Materials for "Addressing challenges in selecting dietary risk factor-outcome pairs for health impact assessments: Developing an evidence database and novel approach to dose-response extraction"

**Appendix A**: Excluded qualified systematic reviews from the NNR2023

**Appendix B:** Food group and dietary risk factor description and mapping - NNR and GBD

**Appendix C**: Search method for the update of the NNR qualified systematic reviews

**Appendix D:** Side by side evidence comparison for conflicting associations

**Appendix E:** Protocol for piecewise non-linear curve extraction

**Appendix F:** GBD Burden of Proof Summary

**Appendix A:** Excluded qualified systematic reviews from the NNR2023

Table A1. Qualified systematic reviews from NNR2023 relating to foods or food groups excluded based on selection criteria.

| First Author | Year | Title | Exposure | Outcome | Commissioned | Reason for Exclusion |
|---|---|---|---|---|---|---|
| Sonestedt et al. (1) | 2012 | Does high sugar consumption exacerbate cardiometabolic risk factors and increase the risk of type 2 diabetes and cardiovascular disease? | Sugar consumption (intrinsic, added, and total sugar) | T2D and CVD | NNR 2012 | No dose-response |
| WHO (2) | 2015 | Guideline: Sugars intake for adults and children | Free sugars | Various | WHO | No dose-response |
| Te Morenga et al.* (3) | 2013 | Dietary sugars and body weight: systematic review and meta-analyses of randomised controlled trials and cohort studies | Sugar consumption | Body weight | WHO (2015) | No dose-response |
| Moynihan et al.* (4) | 2014 | Effect on Caries of Restricting Sugars Intake | Sugar consumption | Dental caries | WHO (2015) | No dose-response |
| Mayer-Davis et al. (5) | 2020 | Added Sugars Consumption and Risk of Cardiovascular Disease: A Systematic Review | Sweets | CVD | DGAC2020 | No dose-response |
| Mayer-Davis et al. (6) | 2020 | Beverage consumption and growth, size, body composition, and risk of overweight and obesity: a systematic review | Beverage consumption | Various | DGAC2020 | No dose-response |
| EFSA (7) | 2022 | Tolerable upper intake level for dietary sugars | Free sugars | Tolerable upper limit | EFSA | No dose-response |
| Rios-Leyvraz & Montez (8) | 2022 | Health effects of the use of non-sugar sweeteners: a systematic review | Non-sugar sweeteners | Various | WHO | No dose-response |
| Rousham et al. (9) | 2022 | Unhealthy food and beverage consumption in children and risk of overweight and obesity: A systematic review and meta-analysis | Sweets | Overweight, obesity | WHO | No dose-response |

| | | | | | | |
|---|---|---|---|---|---|---|
| Fogelholm et al. (10) | 2012 | Dietary macronutrients and food consumption as determinants of long-term weight change in adult populations: a systematic literature review | Macronutrients from various sources | Weight change | NNR2012 | No dose-response |
| Åkesson et al. (11) | 2013 | Health effects associated with foods characteristic of the Nordic diet: a systematic literature review | Potatoes, berries, milk and milk products, red meat | CVD, T2D, inflammatory factors, cancers, bone health, iron status | NNR2012 | No dose-response |
| Lamberg-Allardt et al. (12) | 2023 | Animal versus plant-based protein and risk of cardiovascular disease and type 2 diabetes: a systematic review of randomized controlled trials and prospective cohort studies | Plant protein | CVD and T2D | NNR2023 | No dose-response |
| Snetselaar et al. (13) | 2020 | Seafood consumption during pregnancy and lactation and neurocognitive development in the child: a systematic review | Seafood | Neurodevelopment | DGAC2020 | No dose-response |
| Snetselaar et al. (14) | 2020 | Seafood consumption during childhood and adolescence and cardiovascular disease: A systematic review | Seafood | CVD | DGAC2020 | No dose-response |
| Norwegian Scientific Committee for Food and Environment (15) | 2022 | Benefit and risk assessment of fish in the Norwegian diet | Fish | Various | Norwegian Scientific Committee for Food and Environment | Secondary investigation of dose-response |
| Ramel et al. (16) | 2023 | White meat consumption and risk of cardiovascular disease and type 2 diabetes: a systematic review and meta-analysis | White meat | T2D and CVD | NNR2023 | No dose-response |
| Mayer-Davis et al. (17) | 2020 | Alcohol Consumption and All-Cause Mortality: A Systematic Review | Alcoholic drinks | All-cause mortality | DGAC2020 | No dose-response |

| | | | | | | |
|---|---|---|---|---|---|---|
| CCSUA (18) | 2023 | Canada's Guidance on Alcohol and Health: Final Report | Alcoholic drinks | Various | Health Canada | No dose-response |

*Not qualified systematic reviews but direct underlying evidence for WHO 2015 Report on free sugars. CVD = cardiovascular diseases, T2D = type 2 diabetes.

**Appendix B**: Food group and dietary risk factor description and mapping - NNR and GBD

Table B1. Visual mapping between food groups defined by NNR, food items or groups found in NNR related reviews, and GBD dietary risk factors. The food groups and dietary risk factors colored in orange are common to both frameworks.

| NNR Food Groups | Data from NNR systematic reviews & update | GBD dietary risk factors |
|---|---|---|
| Beverages | Coffee | |
| | Tea | |
| | Mate | |
| | SSBs | SSBs |
| | ASBs | |
| Cereals | Whole grain | Whole grain |
| | Dietary fibre | Fibre |
| | Insoluble fibre | |
| | Vegetable fibre | |
| | Legume fibre | |
| | Soluble fibre | |
| Vegetables, Fruits & Berries | Vegetable | Vegetables |
| | Fruit | Fruit |
| | Fruits & Vegetable | |
| | Preserved vegetables | |
| Potatoes | | |
| Fruit juices | | |
| Pulses/Legumes | | Legumes |
| Nuts & Seeds | | Nuts & Seeds |
| Fish & seafood | | Seafood Omega-3 Fatty Acid |
| Red meat | Total protein | |
| | Animal protein | |
| | Total meat | |
| | Processed meat | Processed meat |
| | Red meat | Red meat |
| White meat | | |
| Milk & dairy products | | Milk |
| Eggs | | |
| Fats & Oils | Olive Oil | |
| | Alpha Linoleic Acid | |
| | Linoleic Acid | |
| | LC Omega 3 Fatty Acid | Seafood Omega 3 Fatty Acid |
| | Trans Fatty Acid | Trans Fatty Acid |
| Sweets | | |
| Alcohol | | *Alcohol** |
| | | PUFA |
| | | Calcium |

*Not a dietary risk factor in GBD but still a reported risk factor, included for comparison in Appendix F.

**Detailed definitions of the food groups are taken verbatim from supporting GBD information and the NNR2023 main document** (19,20)**.**

Beverages

- **NNR**: No definition (water, coffee, tea, SSBs, ASBs)
- **GBD SSBs drinks**: Beverages with ≥50 kcal per 226.8mL serving, including carbonated beverages, sodas, energy drinks, fruit drinks, but excluding 100% fruit and vegetable juices

Fibres

- **GBD fibre:** From all sources including fruits, vegetables, grains, legumes, and pulses

Cereals

- **NNR**: The definition of cereals (grains) comprises commonly eaten seeds from species from the grass family, i.e. wheat, rye, oat, barley, maize, rice, millet, sorghum/durra, teff and wild rice (21). In addition, the global consensus definition includes 'pseudo-cereals' (amaranth, buckwheat and quinoa) (22). Whole grains are defined as intact grains or processed grains (e.g. ground, cracked or flaked) where the three fractions endosperm, germ and bran are present in the same relative proportion as in the intact grains (22). A consensus statement suggests that whole grain should be the main ingredient in whole grain food products, i.e. whole grain should constitute more than 50% of the dry matter (23). The term "cereals" also encompasses refined grains, where the refining process involves removing the bran and germ, which are nutrient-rich components, leaving the starchy endosperm, containing varying amounts of protein (e.g. gluten). Many whole grain breads also contain refined grains for taste and baking properties.
- **GBD whole grain**: Bran, germ, and endosperm in their natural proportion) from breakfast cereals, bread, rice, pasta, biscuits, muffins, tortillas, pancakes, and other sources

Vegetables, fruits, and berries:

- **NNR**: Products within this food group are culinary defined as vegetables, fruits and berries. Potatoes and pulses are not included as vegetables in the NNR2023 report. Green beans and peas may be included in the vegetable food group**. Fruit juices** derived from fruits and berries also constitute **a separate food group**. Vegetables include cruciferous vegetables, leafy green vegetables, yellow/orange/red vegetables, allium vegetables, and non-starchy root vegetables, such as carrots, beets, parsnips, turnips, and swedes. Fruit subgroups are citrus fruits (e.g. oranges, lemon, lime, grapefruit), stone fruits (e.g. cherries, plums) and pome fruit (e.g. apples, pears). Vegetables, fruits and berries are commonly high in water, low in energy, contain numerous nutrients, and good sources of dietary fibre, vitamin C, vitamin E, vitamin K, folate, and potassium. They also contain other bioactive compounds or phytochemicals, and the synergistic effects of these are still not fully understood. Cruciferous vegetables (Brassica), including broccoli, Brussels sprouts, cabbage, cauliflower, kale, and turnips, are sources of calcium. Additionally, leafy green vegetables such as spinach, Swiss chard, and lettuce offer iron, zinc, calcium, magnesium, and carotenoids, with dark green vegetables particularly rich in carotenoids. Berries are small, juicy and pulpy fruits (24).

- **GBD Fruits**: fresh, frozen, cooked, canned, or dried fruits, excluding fruit juices and salted or pickled fruits
- **GBD Vegetables**: fresh, frozen, cooked, canned, or dried vegetables, excluding legumes and salted or pickled vegetables, juices, nuts, seeds, and starchy vegetables such as **potatoes** or corn

Potatoes

- **NNR**: Potatoes (Solanum tuberosum) is a commonly consumed staple food. Potatoes are not included in the vegetable food group, due to their high content of starch.

Fruit juices

- **NNR**: Fruit juice is 100% pure juice made from whole or flesh of fruits and berries. It is not permitted to add sugar, sweeteners, preservatives, flavouring or colouring to fruit juice (25).

Pulses/legumes

- **NNR**: Pulses are often used as the term for the ripened (or dried) form of peas and beans, including lentils, but **excluding green beans and green peas**.
- **GBD**: Legumes: (fresh, frozen, cooked, canned, or dried legumes)

Nuts & seeds

- **NNR**: A culinary definition of nuts includes **tree nuts, peanuts, and seeds**. Peanuts, almonds, walnuts, hazelnuts, cashews, Brazil nuts, macadamias, pistachios, sesame, and sunflower seeds are some of the frequently consumed nuts and seeds (26).
- **GBD**: nuts and seeds (no definition)

Fish & seafood

- **NNR**: no definition
- **GBD seafood omega-3 fatty acids**: eicosapentaenoic acid and docosahexaenoic acid

Red meat

- **NNR**: Pigs, cattle, sheep, goats, game, moose, deer, ... (includes processed and non-processed meat, sometimes differentiated, sometimes not)
- **GBD red meat**: beef, pork, lamb, and goat, but excluding poultry, fish, eggs, and all processed meats
- **GBD processed meat**: meat preserved by smoking, curing, salting, or addition of chemical preservatives

White meat

- **NNR**: chicken, hen, turkey, duck
- **GBD**: not in the dietary risks factor

Milk & dairy products

- **NNR**: no definition (milk, yoghurt, cheese)
- **GBD milk**: including non-fat, low-fat, and full-fat milk, excluding soy milk and other plant derivatives

Eggs

- **NNR**: chicken eggs

Fats & oils

- **NNR**: no definition (vegetable oils, margarine, butter, butter mixes, shortenings)
- **GDB polyunsaturated fatty acids**: omega-6 fatty acids from all sources, mainly liquid vegetable oils, including soybean oil, corn oil, and safflower oil
- **GDB trans fatty acids**: trans-fat from all sources, mainly partially hydrogenated vegetable oils and ruminant products

Sweets

- **NNR**: no definition (chocolate and other sugary foods such as cakes, biscuits, other confectioneries and **SSBs**)

Alcohol

- **NNR**: no definition (Ethanol in beer, wine and spirit)

**Appendix C**: Search method for the update of the NNR qualified systematic reviews

Table C1. Search methods and results for the updated search based on NNR2023 scoping review background paper methodology for respective food groups. Search conducted August 2nd, 2024.

| Food Group | Search String | Database | Results |
|---|---|---|---|
| ***Beverages*** | Hand selection of articles and sources was used in the original scoping review. | | |
| *Artificially Sweetened Beverages* | artificially sweetened beverages OR low-calorie sweetened AND systematic review AND 2023/04/15: 3000/12/31[Date - Publication] | PubMed | 12 |
| *Coffee and Tea** | coffee OR caffeine OR tea AND systematic review AND 2023/04/15: 3000/12/31[Date - Publication] | PubMed | 188 |
| *Coffee and Tea Mendelian* | Mendelian randomization AND coffee AND 2023/04/15: 3000/12/31[Date - Publication] | PubMed | 85 |
| ***Cereals*** | ((cereal*[Title] OR grain*[Title] OR "whole grain"[Title] OR "whole grains"[Title] OR Edible Grain[MeSH Terms] OR whole grains[MeSH Terms]) AND humans[Filter] AND (systematic review[Publication Type] OR meta-analysis[Publication Type])) AND ((2023/04/15:3000/12/31[Date - Publication])) | PubMed | 13 |
| ***Vegetables, Fruits, and Berries*** | (fruit*[Title/Abstract] OR vegetable*[Title/Abstract] OR berry[Title/Abstract] OR berries[Title/Abstract] OR potato*[Title/Abstract] OR "Fruit"[MeSH Terms:noexp] OR "Vegetables"[MeSH Terms:noexp] OR "Fruit and Vegetable Juices"[MeSH Terms]) AND ("meta analysis"[Publication Type] OR "systematic review"[Publication Type]) AND 2023/04/15:3000/12/31[Date – Publication] | PubMed | 290 |
| *Potatoes (in addition to Vegetables, Fruits, and Berries)* | potato*[Title/Abstract] AND cancer[Title/Abstract] AND 2023/04/15:3000/12/31[Date - Publication] AND ("meta analysis"[Publication Type] OR "systematic review"[Publication Type]) | PubMed | 3 |
| *Fruit Juice (in addition to Vegetables, Fruits, and Berries)* | “fruit juice”[Title/Abstract] AND cancer[Title/Abstract] AND 2023/04/15:3000/12/31[Date - Publication] AND (“meta- | PubMed | 2 |

| | | | |
|---|---|---|---|
| | analysis"[Publication Type] OR "systematic review"[Publication Type]) | | |
| ***Nuts and Seeds*** | De novo systematic review conducted by Årnesen et al. An umbrella review was also conducted in 2022 by Balakrishna et al. No search method available. | | |
| ***Fish and Seafood*** | (fish*[Title/Abstract] OR seafood*[Title/Abstract] OR fish oil*[Title/Abstract]) AND "humans"[MeSH Terms] AND ("meta analysis"[Publication Type] OR "systematic review"[Publication Type]) AND "english"[Language] AND 2023/04/15:3000/12/31[Date - Publication] | PubMed | 138 |
| ***Meat*** | (meat[MeSH Terms] OR meats[MeSH Terms]) AND (2023/04/15:3000/12/31[Date - Publication]) AND humans[Filter] AND (systematic review[Publication Type] OR meta-analysis[Publication Type]) | PubMed | 28 |
| ***Meat*** | ((((ALL=(meat OR meats OR beef OR lamb OR mutton OR pork OR poultry)) AND ALL=(systematic review OR meta-analysis)) AND DT=(Review)) AND WC=(Respiratory System OR Allergy OR Gerontology OR Integrative & Complementary Medicine OR Geriatrics & Gerontology OR Pediatrics OR Behavioral Sciences OR Obstetrics & Gynecology OR Clinical Neurology OR Neurosciences OR Rheumatology OR Hematology OR Peripheral Vascular Disease OR Immunology OR Orthopedics OR Medicine, Research & Experimental OR Surgery OR Psychiatry OR Cardiac & Cardiovascular Systems OR Gastroenterology & Hepatology OR Endocrinology & Metabolism OR Oncology OR Medicine, General & Internal OR Nutrition & Dietetics)) AND DOP=(2023-04-15/2500-12-31) | Web of Science | 95 |
| ***Dairy*** *(Mendelian Randomisation)* | "Mendelian randomization"[Title/Abstract] AND (LCT[Title/Abstract] OR lactase[Title/Abstract] OR milk[Title/Abstract]) AND 2023/04/15:3000/12/31[Date – Publication] | PubMed | 16 |

| | | | |
|---|---|---|---|
| ***Dairy*** | ((milk [MeSH Terms] OR dairy products [MeSH Terms] OR cheese [MeSH Terms]) AND humans[Filter] AND (systematic review[Publication Type] OR meta-analysis[Publication Type])) AND ((2023/04/15: 3000/12/31[Date - Publication])) | PubMed | 94 |
| ***Pulses*** | (pulses[MeSH Terms] OR legumes[MeSH Terms] OR "food groups") AND (cancer OR Neoplasms[MeSH Terms] OR obesity[MeSH Terms] OR dementia[MeSH Terms]) AND ("meta analysis"[Publication Type] OR "systematic review"[Filter]) AND 2023/04/15: 3000/12/31[Date - Publication] | PubMed | 11 |
| ***Eggs*** | (eggs[MeSH Terms] OR egg) AND 2023/04/15: 3000/12/31[Date - Publication] AND "humans"[filter] AND ("review"[Publication Type] OR "systematic review"[Filter] OR "meta-analysis"[Publication Type]) | PubMed | 241 |
| ***Fats and Oils*** | ("dietary fat"[Title/Abstract] AND odds ratio OR "dietary fats"[Title/Abstract] OR "vegetable oil"[Title/Abstract] OR "butter"[Title/Abstract] OR "ghee"[Title/Abstract] OR "corn oil"[Title/Abstract] OR "cottonseed oil"[Title/Abstract] OR "canola"[Title/Abstract] OR "olive oil"[Title/Abstract] OR "rapeseed oil"[Title/Abstract] OR "safflower oil"[Title/Abstract] OR "sunflower oil"[Title/Abstract] OR "sesame oil"[Title/Abstract] OR "soybean oil"[Title/Abstract] OR "plant oil"[Title/Abstract] OR "seed oil"[Title/Abstract] OR "cooking oil"[Title/Abstract] OR "margarine"[Title/Abstract] OR "flaxseed oil"[Title/Abstract] OR "palm oil"[Title/Abstract] OR "coconut oil"[Title/Abstract] OR "camelina oil"[Title/Abstract] OR "lard"[Title/Abstract] OR "mayonnaise"[Title/Abstract] OR "dietary fats"[MeSH Terms] OR "plant oils"[MeSH Terms]) AND ("meta analysis"[Publication Type] OR "systematic review"[Filter]) AND | PubMed | 129 |

| | | | |
|---|---|---|---|
| | "humans"[Filter] AND 2023/04/15: 3000/12/31[Date - Publication] | | |
| ***Sweets*** | (sugary[Title/Abstract] OR candy[Title/Abstract] OR sweets[Title/Abstract] OR chocolate[Title/Abstract] OR sugar-sweetened[Title/Abstract] OR confection*[Title/Abstract] OR biscuit*[Title/Abstract] OR cookie*[Title/Abstract] OR pastr*[Title/Abstract] OR bakery[Title/Abstract] OR cake*[Title/Abstract] OR "ice cream"[Title/Abstract]) AND Humans[Filter] AND Review[Publication Type] AND 2023/04/15: 3000/12/31[Date – Publication] | PubMed | 118 |
| ***Alcohol*** | (Alcoholics[Majr] OR Alcoholic Beverages[Majr] OR Alcohol Drinking[Majr] OR "Alcohol-Related Disorders"[Majr:NoExp] OR "Alcohol-Induced Disorders"[Majr] OR "Alcohols/adverse effects"[Majr:NoExp] OR "Ethanol/adverse effects"[Majr:NoExp] OR "Alcohols/toxicity"[Majr:NoExp] OR "Ethanol/toxicity"[Majr:NoExp] OR ((alcohol[ti] OR alcoholism[ti] OR alcoholics[ti] OR (alcoholic[ti] NOT non-alcoholic[ti]) OR (drink*[ti] NOT water[ti]) OR drunk*[ti] OR beer*[ti] OR wine[ti] OR wines[ti] OR (ethanol[ti] AND (consum*[ti] OR drink*[ti] OR drunk*[ti] OR intake[ti] OR intox*[ti] OR abus*[ti] OR misuse[ti] OR mis-use[ti]))) NOT medline[sb])) AND ("Systematic Review"[Publication Type] OR "Meta-Analysis"[Publication Type] OR metaanalysis[Title] OR metaanalyses[Title] OR "meta-analysis"[Title] OR "meta-analyses"[Title] OR (systematic[Title] AND review[Title]) OR "synthesis review"[Title] OR metasynthesis[Title] OR metasyntheses[Title] OR "meta-synthesis"[Title] OR "meta-syntheses"[Title] OR metaregression[Title] OR "meta-regression"[Title] OR "synthesis of evidence"[Title] OR "evidence synthesis"[Title] OR "evidence syntheses"[Title] OR "evidence-based synthesis"[Title] OR "scoping review"[Title] OR "umbrella review"[Title] OR | PubMed | 160 |

| | |
|---|---|
| | ((review[Title] OR overview[Title] OR summary[Title] OR synthesis[Title] OR syntheses[Title]) AND "systematic reviews"[Title]) OR "Mendelian Randomization Analysis"[Mesh] OR "Mendelian Randomization"[Title]) AND (English[lang] OR Norwegian[lang] OR Swedish[lang] OR Danish[lang]) AND 2023/04/15: 3000/12/31[Date - Publication] NOT "pancreatitis 1"[All Fields] NOT "smoking"[All Fields] NOT "psychiatry"[All Fields] NOT "multiple sclerosis"[All Fields] |

*search was conducted September 5th, 2024.

**Appendix D:** Side by side evidence comparison for conflicting associations.

The studies highlighted in green are the ones selected after the side-by-side comparison.

Table D1. Comparison of evidence sources for whole grain and health outcome colorectal cancer.

| Health Outcome | **CRC** | |
|---|---|---|
| Main Author | Reynolds (27) | WCRF 2018 (28) |
| Association | 0.97 [0.95-0.99]/15g/d | 0.83/90g/d, no evidence of non-linear |
| # of Studies | 8 | 6 |
| Quality of Evidence | Moderate | Probable – decreasing risk |
| Other Measures | I2 = 45% | I2= 18.2, p=0.295 |
| AMSTAR-2 | / | / |
| Studies | Egeberg (2010), Fung (2010), McCullough (2013), Kyrø (2013), Larsson (2005), Pietinen (1999), Schatzkin (2007) (High to Low comparison) | Kyrø (2013), Fung (2010), Schatzkin (2007), McCarl (2006), Larsson (2005) |
| Funding | WHO | WCRF |

Table D2. Comparison of evidence sources for total dietary total fibre and health outcome all-cause mortality.

| Health Outcome | **All-Cause Mortality** | |
|---|---|---|
| Main Author | Mirrafiei (29) | Reynolds (27) |
| Association | 0.85 [0.81-0.88] 10g/day | 0.93[0.90-0.95] 8g/day |
| | HR | RR |
| # of Studies | 14 | 5 |
| Quality of Evidence | Moderate | Moderate |
| Other Measures | inverse parabolic association between total fiber intake and all-cause mortality risk (Pnonlinearity < 0.001, Pdose–response < 0.001) | I2=38%, Phet=0.002 |
| AMSTAR-2 | / | / |
| Studies | Donmingez (2019), Katagiri (2020), Partula (2020), Akbaraly (2011), Bazzano (2003), Buil-Cosiales (2014), Chan (2016), Chuang (2012), Park (2011), Streppel (2008), Todd (1999), Xu (2014), Kwon (2022), Xu (2022) | Buil-Cosiales (2014), Xu (2016), Chan (2016), Bazzano (2003), Chuang (2012) |
| Funding | No funding information | Health Research Council of New Zealand, WHO, Riddet Centre, Healthier Lives National Science Challenge, University of Otago, Otago Southland Diabetes Trust |

Table D3. Comparison of evidence sources for total dietary total fibre and health outcome CVD mortality.

| Health Outcome | CVD Mortality | |
|---|---|---|
| Main Author | Mirrafiei (29) | Reynolds (27) |
| Association | HR | RR |
| # of Studies | 13 | 10 |
| Quality of Evidence | Moderate | Moderate |
| Other Measures | The risk of CVD mortality decreased linearly within total fiber intake of 7.5 to 25 g d−1, but the risk did not change remarkably at higher intake (Pnonlinearity < 0.001, Pdose–response < 0.001) | I2=44%, phet=0.315 |
| AMSTAR-2 | / | / |
| Studies | Katagiri (2020), Miyazawa (2020), Akbaraly (2011), Eshak (2010), Bazzano (2003), Buil-Cosiales (2014), Buyken (2010), Chuang (2012), Park (2011), Threapleton (2008), Xu (2014), Kwon (2022), Xu (2022) | Buil-Cosiales (2014), Buyken (2010), Chaung (2012), Eshak (2010) Supplemental, Park (2011), Threapleton (2013) |
| Funding | No funding information | Health Research Council of New Zealand, WHO, Riddet Centre, Healthier Lives National Science Challenge, University of Otago, Otago Southland Diabetes Trust |

Table D4. Comparison of evidence sources for total dietary total fibre and health outcome cancer mortality.

| Health Outcome | **Cancer Mortality** | |
|---|---|---|
| Main Author | Mirrafiei (29) | Reynolds (27) |
| Association | 0.90 [0.84-0.96] 10g/day | 0.94[0.92-0.96] 8g/day |
| | HR | RR |
| # of Studies | 7 | 5 |
| Quality of Evidence | Moderate | Moderate |
| Other Measures | modest inverse association between dietary fiber intake and cancer mortality risk (Pnonlinearity = 0.11, Pdose–response = 0.01) | I2=20%, Phet=0.837 |
| AMSTAR-2 | / | / |
| Studies | Katagiri (2020), Buil-Cosiales (2014), Chan and Lee (2016), Chuang (2012), Park (2011), Xu (2014), Xu (2022) | Buil-Cosiales (2014), Chuang (2012), Xu (2014), Xu (2016), Chan (2016) |
| Funding | no funding info | Health Research Council of New Zealand, WHO, Riddet Centre, Healthier Lives National Science Challenge, University of Otago, Otago Southland Diabetes Trust |

Table D5. Comparison of evidence sources for tea and health outcome bladder cancer.

| Health Outcome | **Bladder Cancer** | |
|---|---|---|
| Main Author | WCRF 2018 (30) | Qi 2023 (31) |
| Association | 0.94[0.89-0.98] 1cup/day | 0.97[0.94-0.99] 1cup/day |
| | RR | RR |
| # of Studies | 4 | 6 |
| Quality of Evidence | Limited - Suggestive | Moderate |
| Other Measures | I2=0%, pheterogeneity = 0.42 | I2=12.3%, |
| AMSTAR-2 | | |
| Studies | Ros M (2011), Tripathi (2002), Zeegers (2001), Michaud (1999) | Hashmian (2019), Chyou (1993), Heilbrun (1986), Kurahashi (2009), Michaud (1999), Nagano (2000) |
| Funding | WCRF | Beijing Advanced Innovation Center for Food Nutrition and Human Health, the National Natural Science Foundation of China, the Chinese Universities Scientific Fund, and the Beijing Municipal Natural Science Foundation |

**Appendix E:** Protocol for Piecewise Non-Linear Curve Extraction

Following the first two steps (pure extraction of points from source and contacting respective authors for meta-analysis results), the final option for obtaining an estimate of the non-linear dose-response data required was to do a self-extraction. The following protocol outlines the detailed steps involved in this process.

**Software:** RStudio (version 4.4.1), WebPlotDigitiser V5.2 (32,33)

**Steps:**

1. Store an image of the plot of interest. If it is not available directly from the source as an image, the user may need to use a screen capture tool on their device. Note: if there is a legend or other imagery on the plot, this should be removed from the image as the automatic extraction tool relies on colours in the plot.
2. Upload the image to WebPlotDigitiser, follow the provided instructions to calibrate the axes of the plot with the tool. Complete the process by entering the axes values and clicking calibrate on the right panel.
3. Using the "Automatic Extraction" option in the right panel: select the relevant colour from dominant colors options for the curve of interest. For "Algorithm", select "X Step w/ Interpolation". Some small calculations may be needed to be done manually here. For standardisation, the goal is to have 50 points plotted on the curve, however due to curve drawing and variation, this may not always be the case.

    X_max – X_min / 50 = change in X step
    e.g. Whole Grains:
    Plot ends at 83g
    83-0/50 = 1.66

4. Visually inspect the data points and ensure they follow the curve of the desired plot.
5. "View Data" and download the CSV file.
6. Load the CSV into RStudio using the provided RScript.
7. Run the script to create a piecewise function from the points, this function can be used in projects, etc.

**Appendix F:** GBD Burden of Proof Summary

Table F1. Burden of Proof Summaries for food-based dietary risk factors (34). Only associations with two or more stars or more were included. Associations already included in the NNR2023 or the updated search were not included in this table.

| Risk Factor | Exposure | Outcome | Mean RR for the Exposure | Exposure range (g/day) | Strength of the relationship | Risk-Outcome Score (ROS) |
|---|---|---|---|---|---|---|
| High processed meat | Increasing intake of processed meat | T2D | 1.22 | 0-51.7 | 2 stars | 0.075 |
| | | IHD | 1.02 | 0-38.6 | 2 stars | 0.011 |
| | | CRC | 1.09 | 0-38.51 | 2 stars | 0.023 |
| Low fruits | Increasing intake of fruits | Tracheal, bronchus, and lung cancer | 0.73 | 15.6-368.6 | 3 stars | 0.19 |
| | | IHD | 0.84 | 22.6-451.4 | 2 stars | 0.062 |
| | | Ischemic stroke | 0.79 | 8.3-496.8 | 3 stars | 0.15 |
| | | Hemorrhagic stroke | 0.84 | 8.3-491.4 | 2 stars | 0.029 |
| | | T2D | 0.90 | 12.4-329.7 | 2 stars | 0.031 |
| Low whole grains | Increasing intake of whole grains | CRC | 0.88 | 9.9-230.3 | 2 stars | 0.07 |
| | | IHD | 0.78 | 3.3-197.6 | 3 stars | 0.78 |
| | | T2D | 0.79 | 2.3-250.7 | 2 stars | 0.094 |
| Low nuts and seeds | Increasing intake of nuts and seeds | IHD | 0.79 | 0-23.5g | 2 stars | 0.11 |
| Low in milk | Increasing intake of milk | CRC | 0.86 | 0-656.6 | 2 stars | 0.064 |
| Low fibre | Increasing intake of fibre | CRC | 0.8 | 4.3-26.5 | 2 stars | 0.12 |
| | | IHD | 0.83 | 3.9-25.5 | 2 stars | 0.12 |
| | | Ischemic stroke | 0.92 | 3.9-22.2 | 2 stars | 0.01 |
| | | T2D | 0.84 | 4.4-30.3 | 2 stars | 0.12 |
| Low seafood omega-3 | Increasing intake of seafood omega-3 | IHD | 0.76 | 0.025-1.73 | 2 stars | 0.1 |
| High SSBs | Increasing intake of SSBs | IHD | 1.05 | 0-365 | 2 stars | 0.023 |
| | | T2D | 1.16 | 1.54-389.76 | 2 stars | 0.073 |

| | | | | | | |
|---|---|---|---|---|---|---|
| High alcohol use | Increasing intake of alcohol | IHD | 1.00 | 36.31-45.00 | 2 stars | 0.0011 |
| | | Intercerebral stroke | 1.31 | 5.04-49.97 | 2 stars | 0.049 |
| | | T2D | 1.07 | 9.00-37.50 | 2 stars | 0.037 |

CRC = colorectal cancer, IHD = ischaemic heart disease, T2D = type 2 diabetes.

33. Rohgatgi A. WebPlotDigitizer [Internet]. Available from: https://automeris.io

34. Institute for Health Metrics and Evaluation [Internet]. [cited 2024 Sept 12]. Burden of Proof. Available from: https://vizhub.healthdata.org/burden-of-proof/
